# Cognitive Health Among Middle-aged and Older Adults in India - the Role of Depression

**DOI:** 10.1101/2025.11.12.25340069

**Authors:** Shreyas Nadkarni, Souvik Banerjee, Aishwarya Putta, Amresh Hanchate

## Abstract

Depression is a well-established risk factor for impaired cognitive function, but the mechanisms underlying this relationship remain poorly understood. In this study, we utilized data from the 2017-18 Wave 1 of the Longitudinal Aging Study in India to examine the potentially causal association between depressive symptoms and cognitive performance in middle-aged and older adults using alternative estimation methods to account for observed and unobserved confounding. We also investigated the mediating roles of activities of daily living (ADL), instrumental activities of daily living (IADL), and sleep problems in the pathway linking depressive symptoms to cognitive decline. Our findings indicated significant decline in cognitive function associated with depression, particularly in the domains of language, immediate memory, and orientation. Moreover, we identified substantial mediation effects, with IADL accounting for 26.3%, ADL for 15.5%, and sleep problems for 10.8% of the relationship between depression and cognitive performance. These results suggest that interventions aimed at improving mental health in aging populations could enhance both performance in daily functioning and cognitive outcomes.

## 1. Introduction

Depression is a debilitating condition that affects around 280 million people worldwide (WHO 2023). In addition to the significant functional impairments and reduction in quality of life associated with depression, it has been found that more than 50% people suffering from Major Depressive Disorder (MDD) exhibit cognitive impairment (Liu et al., 2023). Several hypotheses have been proposed to explain the biological pathways through which depression affects cognitive functioning. Zackova et al. (2021) discuss the shared volumetric reductions in particular brain regions such as the insula, associated with socio-emotional processing, sustained attention, executive function, and superior temporal gyrus (STG), a part of the language network. These reductions are associated with deficits in communication activities, social withdrawal, and reduced participation in mentally stimulating activities, which are risk factors for both major depression and mild cognitive impairment. From a neurochemical perspective, depression causes a cascade of glucocorticoids that leads to a reduction in the volume of the hippocampus, an important region associated with memory formation, as well as cerebrovascular disease, which accelerates the onset of dementia and Alzheimer’s disease (AD) (Butters et al., 2008; Sawyer et al., 2012). Disruption of the synthesis and release of Brain-derived Neurotrophic Factor (BDNF) is also considered to underline several psychiatric and neurological disorders (Diniz et al., 2013). Recent evidence has shown the mediating role of neurofilament light-chain (NfL) in the link between depression and cognitive decline (Xu et al., 2024). In short, depression causes changes in an individual’s neurochemistry and brain structure which can manifest in the form of social withdrawal, communication problems, sleep problems, and difficulty in performing regular activities, which in turn increases the odds of cognitive decline.

Depression is found to affect different cognitive domains differently (Rock et al., 2014; Ahern et al., 2024). Depression leads to the most pronounced deficits in areas of information processing speed, visual learning and memory, and verbal learning and memory (Krieshche et al., 2022). The number of depressive episodes is positively correlated with the extent of cognitive decline across multiple domains - global cognitive status, processing speed, auditory attention capacity, visual attention accuracy, memory (both verbal and visual), verbal fluency, and task-shifting abilities (Semkovska et al., 2019).

Empirical evidence on the relationship between depression and cognitive health in the context of low- and middle-income countries (LMICs) is limited. Thibaut et al. (2023) conducted a meta-analysis from 11 LMICs and concluded that depression was significantly associated with cognitive impairment, particularly in the domain of memory. Smith et al. (2022) analyzed studies on depression and subjective cognitive complaints (SCCs; measured on a scale of 0 to 100 with higher scores indicating worse cognitive function) in 47 LMICs with data from 237,952 individuals aged 18 and above and found significant associations; compared to the group having no depression, those with subsyndromal depression, brief depressive episodes, and depressive episodes had higher mean SCC scores. Muhammad and Meher (2021) used the Longitudinal Ageing Study in India (LASI) Wave 1 data from 2017-18 to estimate the prevalence and correlates of depression and cognitive impairment as well as the association between late-life depression and cognitive health among the older adult population (60+ years) in India. The study noted that individuals suffering from depression were 22% more likely to have cognitive impairment compared to those without depression. In a related study, using the same dataset, Muhammad (2022) highlighted the role of religious participation and religiosity as a significant protective factor in the association between depressive symptoms and cognitive health.

The existing evidence on the relationship between depression and cognitive health cannot be termed as causal as these studies do not account for systematic differences – observed and unobserved – between those with and without depression. Further, self-reports of depressive symptoms can be fraught with errors in measurement if individuals either under-report or over-report their symptoms. In addition, a decline in cognitive health can lead to depression, highlighting the role of a reverse causal pathway. Finally, unobserved personality traits such as extroversion and neuroticism can be correlated with one’s mental health status and with their cognitive health.

In this study, we examine the causal effect of depression on cognitive functioning using alternative linear regression methods to account for observed and unobserved confounding. We use inverse probability of treatment weighting (IPTW), based on propensity scores, to balance the observed characteristics of those with and without depression. We estimate instrumental variable regression models by exploiting a plausibly exogenous variation in the respondent’s depression status by using the spouse’s depression status as an instrumental variable (IV). Using data from the Longitudinal Aging Study in India, we also explore the role of socio-economic and demographic correlates of cognition and depression. Further, we assess the mediating role of sleep problems and limitations in Activities of Daily Living (ADL) and Instrumental Activities of Daily Living (IADL) to better understand the pathways linking depression and cognitive impairment.

## 2. Methods

### 2.1 Data

The Longitudinal Ageing Study in India (LASI) Wave 1 (2017-18) surveyed a nationally representative sample of the Indian population aged 45 years and above and their spouse. The data provides detailed information on socio-economic and demographic characteristics, health, cognition, psychosocial factors, retirement, family structure, among others (Chien et al., 2023). LASI contains data from 73,408 respondents across all the states and union territories of India and is one of the databases from Gateway to Global Ageing (Lee et al., 2019). The sampling was based on the 2011 Census from India and comprised a complex multistage, stratified cluster sampling design. Details about the sampling strategy can be found in Chien et al. (2023). We use Version A.3 of the Harmonized LASI, a respondent-level file containing data from 73,408 respondents (Chien et al., 2023).

Our study sample included individuals 50 years and above (20,242 [27.6%] observations dropped) with non-missing values of the depression score (1,508 [2.1%] observations dropped), yielding an analytic sample of 51,658 individuals (24,216 men and 27,442 women). The sample selection flowchart is depicted in Appendix Figure A1.

#### 2.1.1 Cognition

Respondents were assessed on various domains of cognitive performance, including orientation, verbal skills, language, memory, executive functioning, and numerical abilities (Harvey, 2019). We examined an overall measure of cognitive function as well as cognitive performance in different domains. The measure of overall cognition is a factor score generated using a graded item response theory model by Muthen and Muthen (2017) in the harmonized version of the LASI data (Chien et al., 2023). The factor score was converted to a standardized unit with mean=0 and standard deviation=1 (see Appendix Table A1 for a list of indicators used to generate the factor score). In an alternative specification, we defined a dichotomous indicator (0/1) identifying those in the lowest quartile of cognition factor score as cognitively impaired.

We examined individual cognitive domains grouped into six categories: 1) orientation: date naming, place naming, 2) immediate memory (or working memory): 10 word immediate recall task, 3) delayed memory: 10 word delayed recall task, 4) language: verbal fluency, reading, writing, object naming, 3-stage task, 5) executive functioning: backward counting, serial 7s, computing, and 6) visuospatial: drawing a clock, and drawing interlocked pentagons (see Appendix Table A2 for the complete list of items comprising each domain). This classification is an adaptation of the 5-category classification of Gross et al. (2020) with the memory domain split into immediate and delayed memory since the effect of depression is expected to vary across these two domains (Kriesche et al., 2022). For each cognitive domain, a standardized score was computed by subtracting the mean and dividing by the standard deviation (SD) to ensure comparability of the scores across different domains.

#### 2.1.2 Depression

The primary independent variable of interest is the depressive symptom status. A 10-item version of the Center for Epidemiologic Studies Depression (CES-D) Scale (Radloff, L. S., 1977) was used in the LASI to determine the depression symptom status. The scale included 7 “negative” symptoms - trouble concentrating, felt depressed, felt tired or low in energy, afraid of something, felt alone, bothered by things that don’t usually bother you, and everything was an effort - and 3 “positive” symptoms - overall satisfied, hopeful about the future, and felt happy. Respondents reported their symptoms during the week preceding the survey. A Likert scale was used to solicit responses regarding the depressive symptoms: 0 if rarely or never (<1 day), 1 if sometimes (1 or 2 days), 2 if often (3 or 4 days), and 3 if most or all of the time (5-7 days). The “positive” symptoms were reverse coded so that the CES-D score ranged from 0 to 30 with higher scores indicating higher depressive symptomatology. A score of 10 or higher classified individuals as “depressed”, while scores below 10 indicated “non-depressed” status (Andresen et al., 1994, Fu et al., 2022). As a robustness check, we used alternative (higher) thresholds of 12 and 15.

#### 2.1.3 Covariates

We included age, gender, education level, region, coupled status, religion, caste, participation in physical activity, chronic conditions, current working status, and socializing as control variables in our regression analysis. These were categorized as follows: (i) age (ordered categorical; 50-59 [reference], 60-69, 70-79, 80 and above), (ii) education (ordered categorical; “never attended school” [reference], “primary/middle school (Class 1-9)”, “secondary school (class 10-12)”, “degree or diploma”), (iii) gender (binary; male [reference], female), region (binary; urban [reference], rural), (iv) coupled status (binary; single [reference] if divorced/separated/widowed/never married, coupled if married/partnered), (v) religion (unordered categorical; hindu [reference], muslim, christian, other), (vi) caste (unordered categorical; general [reference], scheduled caste (SC), scheduled tribe (ST), other backward classes (OBC)), (vii) vigorous physical activity (binary; hardly ever/never [reference], at least once a month), (viii) moderate physical activity (binary; hardly ever/never [reference], at least once a month), (ix) yoga/meditation (binary; hardly ever/never [reference], at least once a month), (x) work status (binary; currently not working [reference], currently working), (xi) socializing (binary; do not participate in social activities even once a year [reference], participate in social activities at least once a year), and (xii) chronic physical health conditions (continuous [0-8]; number of physical chronic conditions from the following list: hypertension, diabetes, cancer, lung disease, heart problems, stroke, arthritis, and high cholesterol).

#### 2.1.4 Mediators

To explore the potential pathways linking depression with cognitive impairment, we identified ADL, IADL, and quality of sleep as mediators. ADL comprised 6 everyday activities including bathing, dressing, eating, getting in/out of bed, walking across the room, and using the toilet. Difficulty in performing each activity was identified as a dichotomous measure (yes/no). IADL included 7 activities: making telephone calls, managing money, taking medications, shopping for groceries, preparing hot meals, finding an address in an unfamiliar place, and working around the house/garden. Each of these activities was encoded as a dichotomous measure (yes/no). Quality of sleep was characterized using the following 4 variables: difficulty falling asleep, feeling unrested during the day, waking up during the night, and waking up too early. Each of these was categorized as an ordered categorical variable taking values 1-3 (1 if frequently (5 or more nights per week); 2 if occasionally (3-4 nights per week); and 3 if rarely or never (0-2 nights per week)).

Each measure in the mediator domains was standardized to have a mean=0 and SD=1. A composite score for each mediator domain - ADL, IADL, and sleep quality - was calculated as the mean of the standardized variables and used as the mediating variable for analysing the effect of ADL, IADL, and quality of sleep in the pathway between depression and cognitive impairment.

#### 2.1.5 Instrumental Variable

Depression can be potentially endogenous in the relationship between depression and cognitive health due to measurement errors in the self-reports of depressive symptoms, and/or unobserved confounders correlated with mental health status and cognitive health, and/or reverse causal pathway whereby cognitive impairments can lead to depression. To mitigate the potential bias resulting from unobserved confounders, we used a plausibly exogenous measure as an instrumental variable (IV) – spousal depressive symptoms. Prior literature has noted that spousal depressive symptoms is a significant risk factor for one’s own depressive symptoms (Aloen et al., 2001, Han et al., 2023, Litzelman and Yabroff, 2015) due to shared familial, environmental, and behavioral experiences (Hopman and Gerstorf, 2009; Tower and Kasl, 1996), which provides credence to the use of the variable as a relevant IV. On the other hand, we argue that spousal depression does not have a direct effect on one’s cognitive health; it can affect one’s cognitive health through its impact on one’s depression status (exogeneity assumption). The validity of the IV may be compromised if, for example, one’s poor cognitive health directly leads to depressive symptoms of the spouse. Therefore, we perform the IV analysis as a robustness exercise and do not make strong claims about the strict exogeneity of the IV.

### 2.2 Statistical Analysis

We reported weighted means (proportions) for continuous (categorical) variables and performed a two-sample t-test (chi-squared test) to test for differences in mean (proportions) by depressive symptom status. Survey weights were used to account for the complex multi-stage random sampling design. All statistical analysis was performed using Stata 18 (Statacorp, 2023).

We estimated the association between depressive symptom status and cognitive function using alternative models to account for differences across individuals by depressive symptom status. We begin with a linear regression of cognitive function on depressive symptom status controlling for observed confounders (covariates), including age, gender, education, region, coupled status, religion, caste, measures of physical activity, number of chronic conditions, and work status. Using the same regression framework, we examine the socio-economic and demographic correlates of depressive symptoms and cognition. Differences between those with and without depressive symptoms may have differential association with the outcome of interest (“selection on covariates”). Given the rich set of covariates available, we evaluate the causal effect of depressive symptoms on cognitive function under the assumption of selection on observed covariates. We obtained the propensity score of having depressive symptoms based on the other covariates (in the linear regression model above) and estimated a linear regression with inverse probability of treatment weighting (IPTW) to estimate the average treatment effect on the treated (ATT) (Stuart, 2010; Chesnaye et al., 2021; Bettega et al., 2024). To further account for unobserved confounding in estimating the potential impact of depression symptom status on cognitive function, we estimated a linear regression model using spousal depressive symptoms as an IV and the 2 stage least squares estimator (2SLS). In the first stage, the endogenous variable (own depressive symptoms), is regressed on the IV (spouse’s depressive symptoms) and the covariates (e.g. age, gender, education, etc.). The first stage F-statistic on the IV is considered as a measure of strength/relevance of the IV (Cragg and Donald, 1993). In the second stage, the outcome variable (cognitive function) is regressed on the estimated value of depressive symptoms from the first stage and the same set of covariates from the first stage regression. Since the outcomes of interest - overall cognitive function score and cognitive domain scores - were standardized (mean=0 and SD=1), we interpret the treatment effect as the effect of having depressive symptoms on cognitive function score in terms of standard deviations.

To estimate the potential role of the mediators in the association between depressive symptoms and cognitive function, we performed a mediation analysis and decomposed the total effect into direct and indirect effects (Hayes, 2017; Vanderweel, 2016). The relationship between the treatment, outcome, and the mediator can be depicted using a path diagram (see Figure 1). The effect of the treatment variable on the mediator is denoted by *a*, the effect of the mediator on the outcome is shown by path *b*, and the direct effect of treatment on the outcome (not explained by the mediator) is indicated by path *c*’. The effect of the treatment on the outcome that is explained through the mediator can be expressed as *ab*.

**Figure 1.**
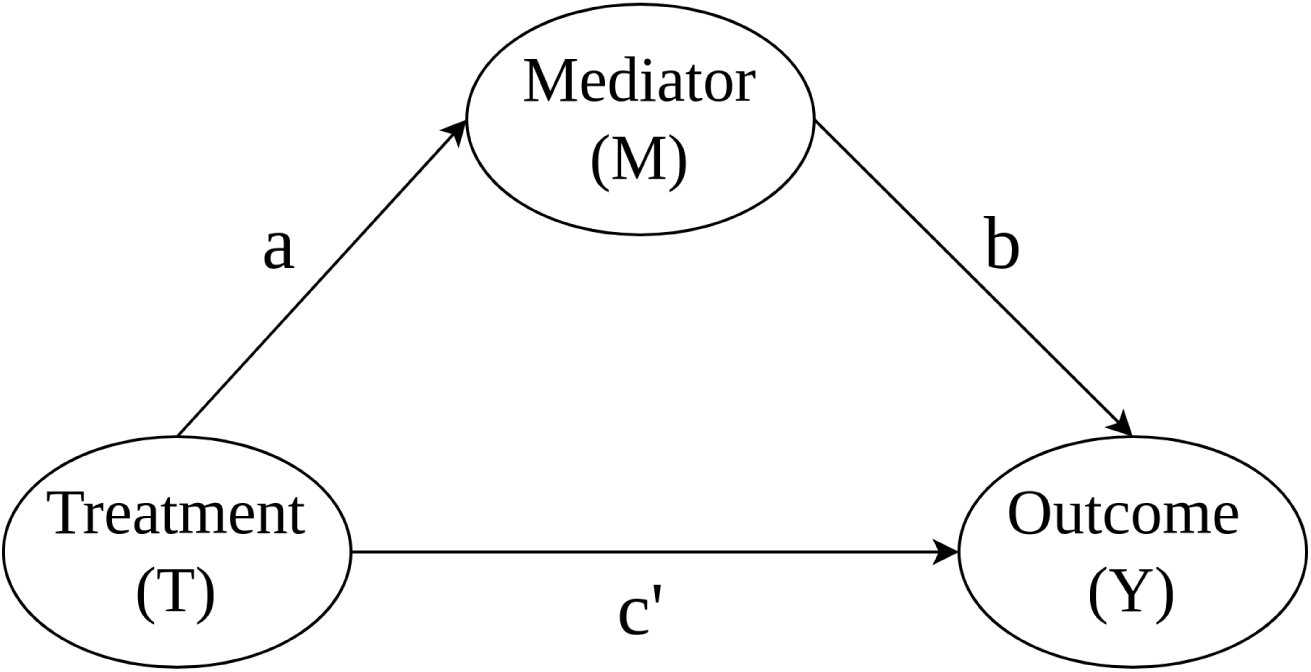
Path diagram for mediation analysis. Notes: The Natural Direct Effect (NDE) is indicated by path *c*’, the Natural Indirect Effect (NIE) is the product of the effect of the Treatment on the Mediator (*a*) and the effect of the Mediator on the Outcome (*b*) and denoted by *ab*. The total effect (TE) of the Treatment on the Outcome is the sum of NDE and NIE (*c*’ + *ab*).

Equivalently, we express the relationship between the outcome variable *Y*, treatment variable *T*, mediator *M*, and a set of covariates *X*, using the following linear regression equations:

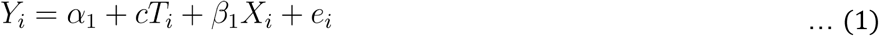

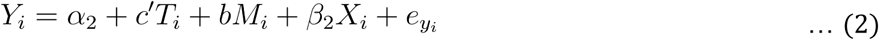

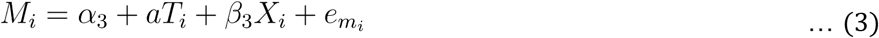

where, *c* denotes the total effect (TE) of *T* on *Y*, controlling for *X* in equation (1). In equation (2), *c*’ is the effect of *T* on *Y* when the effect of *M* on *Y* (denoted by *b*) has been accounted for and is known as the natural direct effect (NDE). In equation (3), *a* denotes the effect of *T* on *M*, controlling for *X*. The natural indirect effect (NIE) is the effect of *T* on *Y* that occurs through *M* and is equal to the product of the effect of *T* on *M* (*a*) and *M* on *Y* (*b*). Therefore,

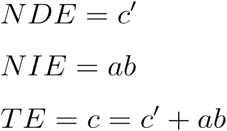

The proportion of the total effect that is mediated through the mediator is defined as *NIE*/*TE* = *ab*/(*c*’ + *ab*).

## 3. Results

### 3.1 Descriptive Statistics

The distributions of the overall cognition factor score and the cognition domain scores are shown in Figure 2, stratified by depressive symptom status. The cognition scores for the depressed category were shifted further left on the distribution, indicating worse cognitive functioning, compared to the non-depressed category.

**Figure 2.**
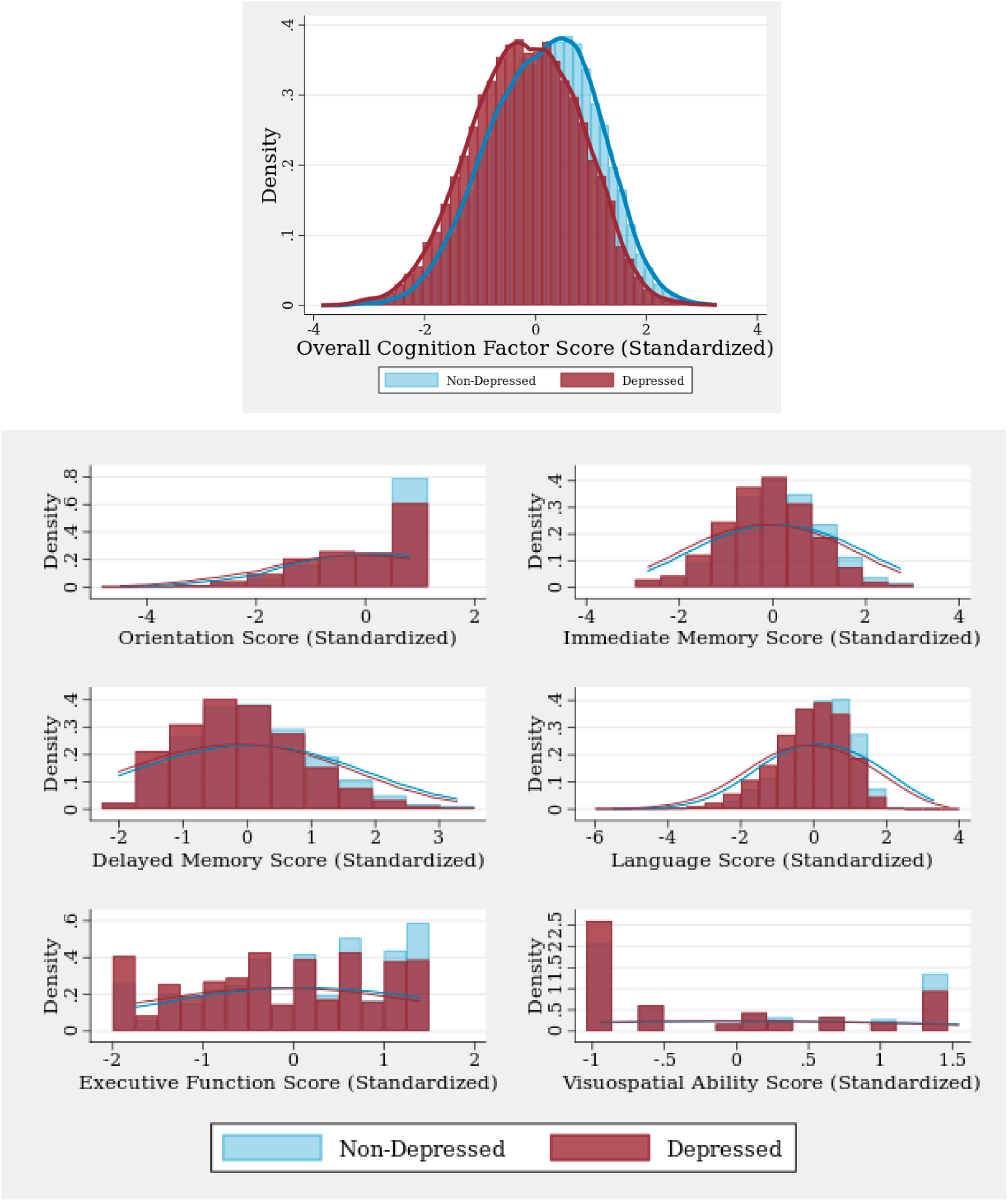
Distribution of Cognition Scores by Depression Status. **Notes**: Higher cognition score indicates better cognitive performance. Each score is standardized to mean = 0 and standard deviation = 1.

Figure 3 shows the summary statistics of the overall cognition score for each of the covariates. We found the strongest gradient in the education categories, with more educated groups having significantly higher cognition scores compared to those who never attended school (p<0.001). We also noted significantly lower cognition scores among the older age groups compared to the 50-60-year age group (p<0.001). The other cohorts associated with significantly worse cognition score included: female; living in a rural area; single status; SC, ST, OBC; Muslim, Christian, Other religion; currently not working; not socializing; and not engaging in physical activities or yoga (all p<0.05).

**Figure 3.**
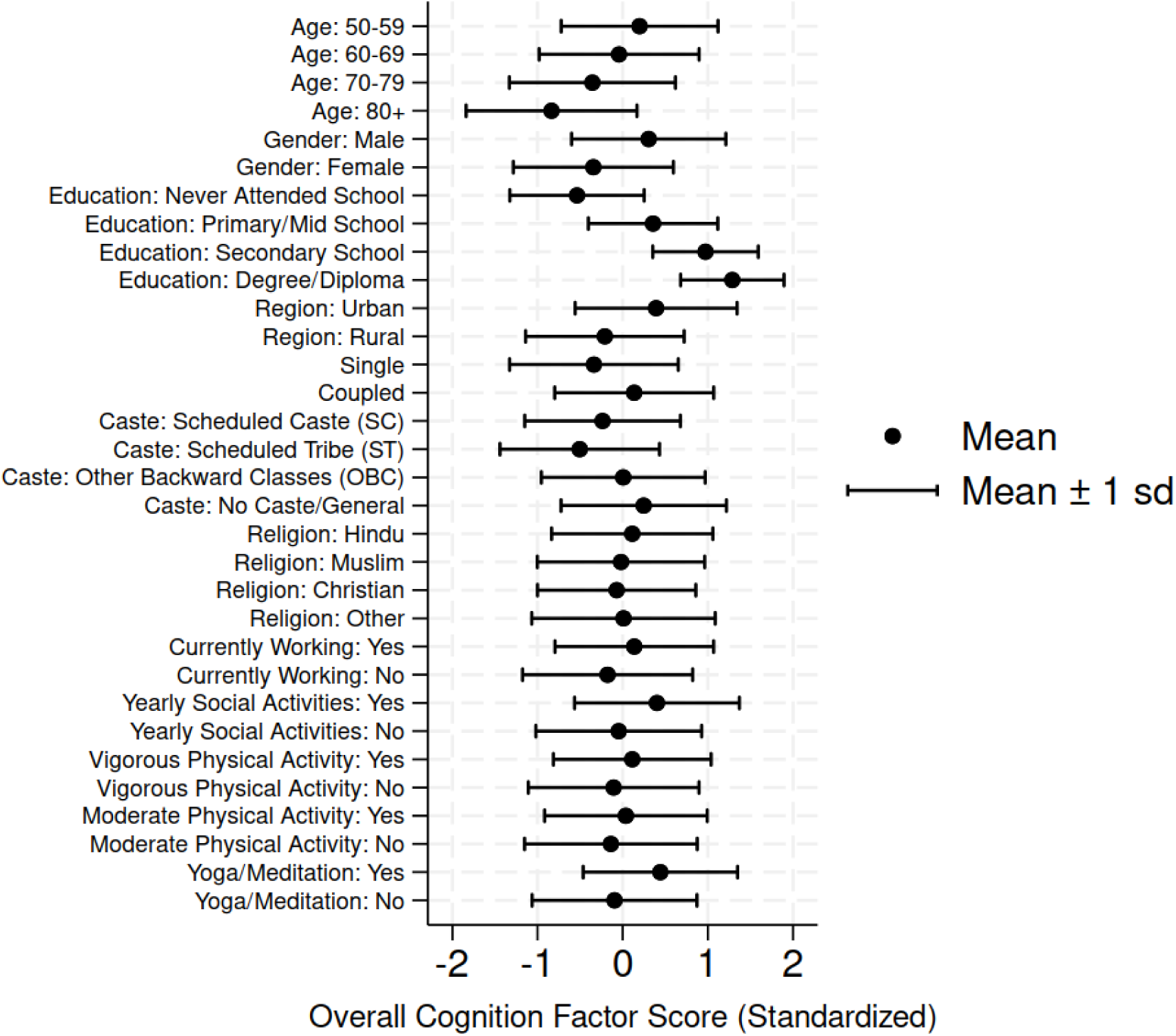
Summary Statistics of Overall Cognition Score. **Notes**: Higher overall cognition score (standardized to mean = 0 and standard deviation = 1) indicates better cognitive performance.

Table 1 shows the descriptive summary statistics of our covariates, and instrumental variable for the full sample as well as stratified by depressive symptom status. The full sample comprised 51,658 individuals, with 22,691 (46.09%) meeting criteria for depressive symptoms. A significantly higher proportion among the depressed group are females, those residing in rural areas, those with single status, those currently not working, compared to the non-depressed group (all p<0.001). Our instrumental variable – spousal depression status – also showed significant association with the respondent’s depression status. Notably, 69.2% of respondents in the depressed group had a spouse with depressive symptoms compared to 24.2% among the non-depressed group (p<0.001). Table 2 shows the descriptive statistics for the overall cognition factor score (standardized), cognition domain scores (standardized), and mediators (standardized). The mean values of the cognition scores were significantly lower for the depressed group compared to the non-depressed group (p<0.001). Similarly, for the mediators, we noted significantly higher mean ADL and IADL scores and significantly lower mean sleep quality score for the depressed group compared to the non-depressed group (all p<0.001). The descriptive statistics of the non-standardized scores are reported in the Appendix (Table A3).

**Table 1.**
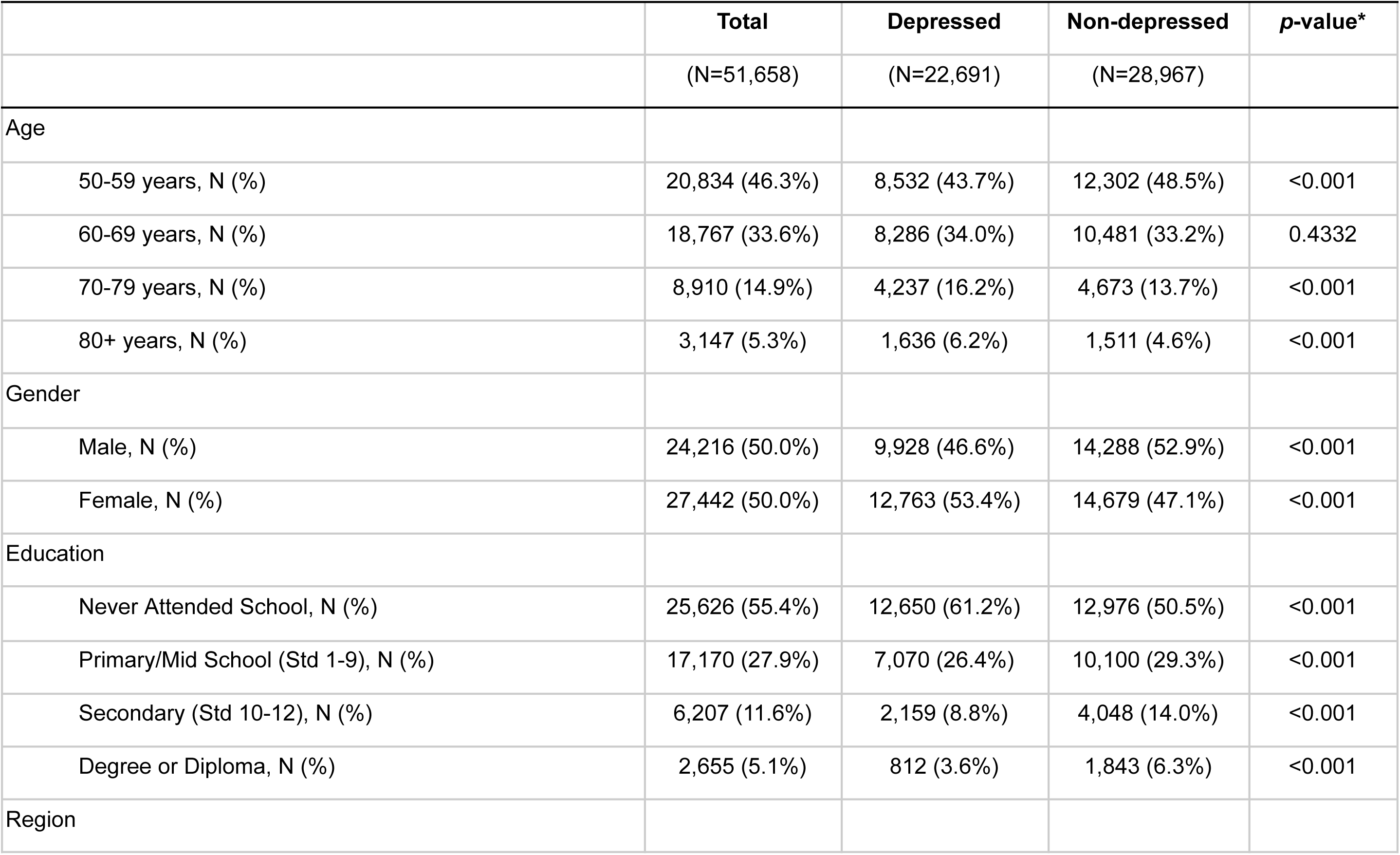
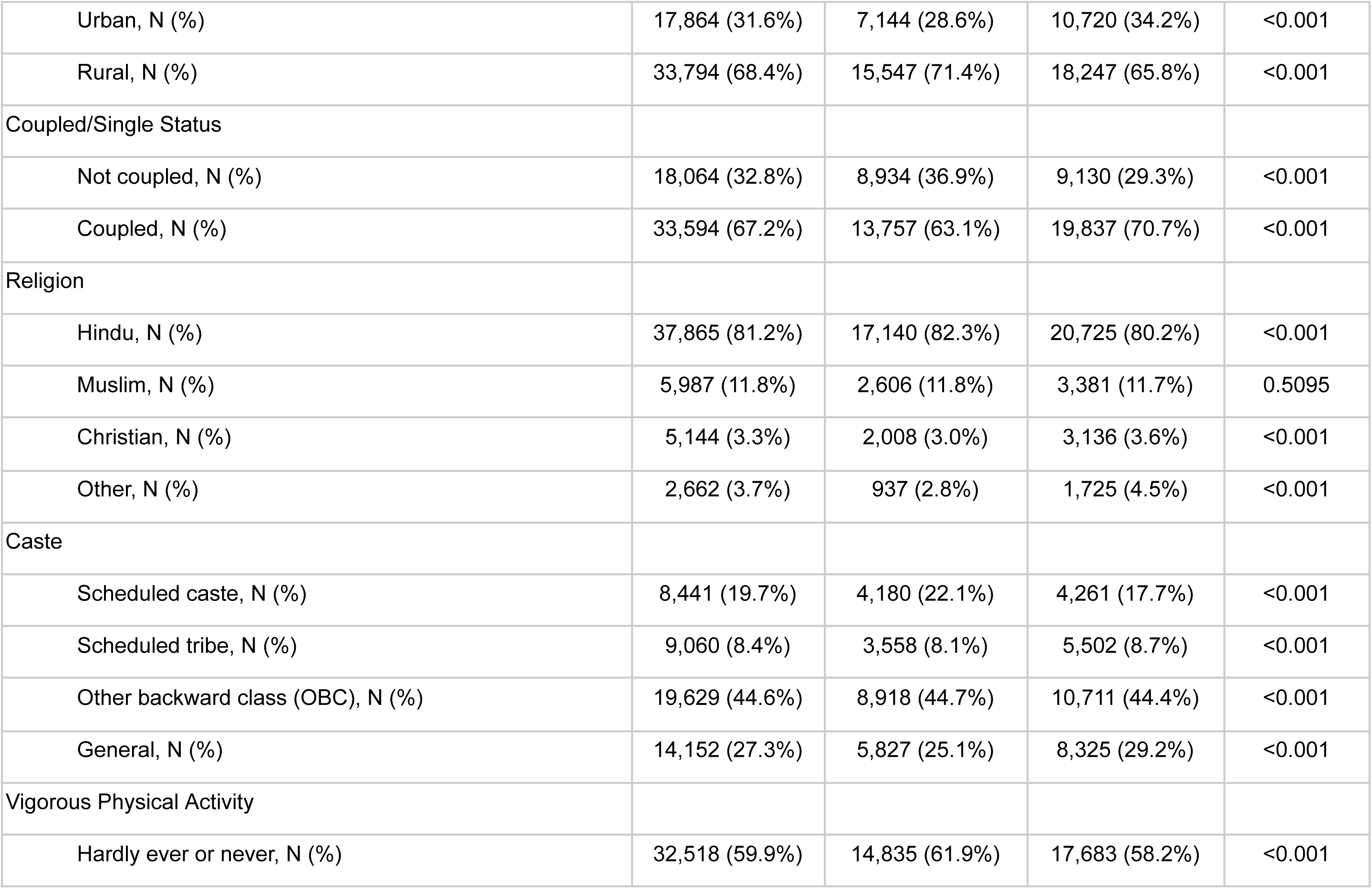
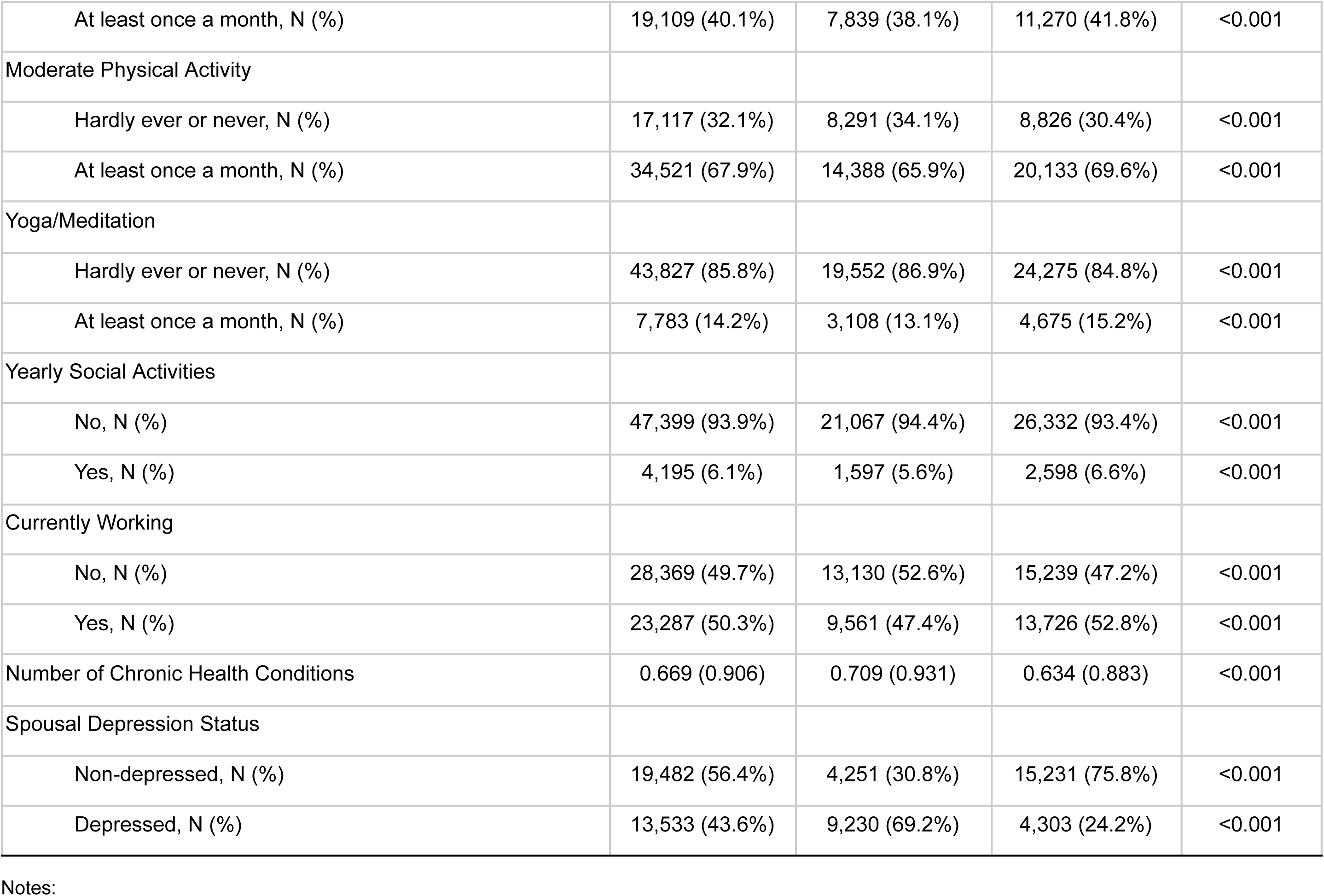
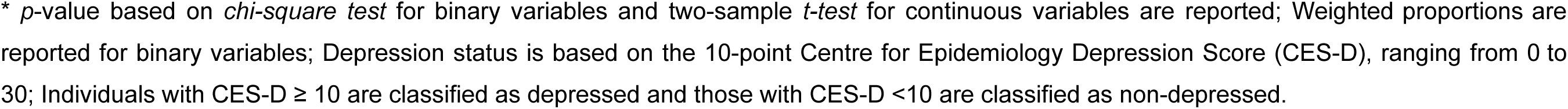
Weighted summary statistics of covariates by depression status.

**Table 2.** Weighted summary statistics for cognition scores and mediators by depression status.

|  | **Total** | **Depressed** | **Non-depressed** | **p-value\*** |
| --- | --- | --- | --- | --- |
|  | (N=51,658) | (N=22,691) | (N=28,967) |  |
| Cognition Scores |  |  |  |  |
| Overall Cognition, Mean (SD)<sup>‡</sup> | -0.018 (0.981) | -0.182 (0.959) | 0.121 (0.977) | <0.001 |
| Orientation, Mean (SD)<sup>‡</sup> | -0.035 (0.983) | -0.165 (1.030) | 0.077 (0.926) | <0.001 |
| Immediate Memory, Mean (SD)<sup>‡</sup> | -0.001 (0.981) | -0.114 (0.957) | 0.095 (0.991) | <0.001 |
| Delayed Memory, Mean (SD)<sup>‡</sup> | 0.002 (0.984) | -0.091 (0.941) | 0.081 (1.014) | <0.001 |
| Language, Mean (SD)<sup>‡</sup> | 0.043 (0.967) | -0.094 (0.990) | 0.134 (0.940) | <0.001 |
| Executive Function, Mean (SD)<sup>‡</sup> | 0.022 (0.990) | -0.100 (0.990) | 0.126 (0.979) | <0.001 |
| Visuospatial Ability, Mean (SD)<sup>‡</sup> | -0.073 (0.978) | -0.179 (0.937) | 0.018 (1.003) | <0.001 |
| Activities of Daily Living (ADL), Mean (SD)<sup>‡</sup> | -0.001 (0.985) | 0.134 (1.164) | -0.116 (0.781) | <0.001 |
| Instrumental Activities of Daily Living (IADL), Mean (SD)<sup>‡</sup> | 0.024 (1.012) | 0.210 (1.125) | -0.134 (0.873) | <0.001 |
| Sleep Quality, Mean (SD)<sup>‡</sup> | -0.032 (1.033) | -0.254 (1.157) | 0.158 (0.870) | <0.001 |

Notes:
\* *p*-value based on two-sample *t*-test for continuous variables are reported; Depression status is based on the 10-point Centre for Epidemiology Depression Score (CES-D), ranging from 0 to 30; Individuals with CES-D $\geq 10$ are classified as depressed and those with CES-D $<10$ are classified as non-depressed.
<sup>‡</sup> Measure standardized to mean 0 and standard deviation 1.

### 3.2 Regression Analysis

Table 3 presents the results of the association of depression status with overall cognition score, controlling for potential confounders. Across all the three regression models, depression status is associated with worse cognition score (OLS: b=-0.112, p<0.001; IPTW: b=-0.115, p<0.001; 2SLS: b=-0.137, p<0.001). We found an age gradient in cognitive function with the older age cohorts having significantly worse cognitive performance: 60-69 years (b=-0.134, p<0.001), 70-79 years (b=-0.322, p<0.001), 80 and above (b=-0.607, p<0.001). Female (b=-0.310, p<0.001) and rural (b=-0.226, p<0.001) respondents had significantly worse cognition scores compared to their male and urban counterparts, respectively. Compared to those who never attended school, the educated groups performed better in terms of cognitive performance: primary/middle school (b=0.664, p<0.001), secondary school (b=1.123, p<0.001), and degree/diploma (b=1.341, p<0.001). Living with a partner (OLS: b=0.109, p<0.001), engaging in moderate physical activity (b=0.106, p<0.001), yoga (b=0.148, p<0.001), and yearly social activities (b=0.088, p<0.001) were significant predictors of higher cognition score. Individuals belonging to scheduled caste (b=-0.083, p<0.001) or scheduled tribe (b=-0.334, p<0.001) had significantly worse cognitive score compared to those from the general caste.

**Table 3.**
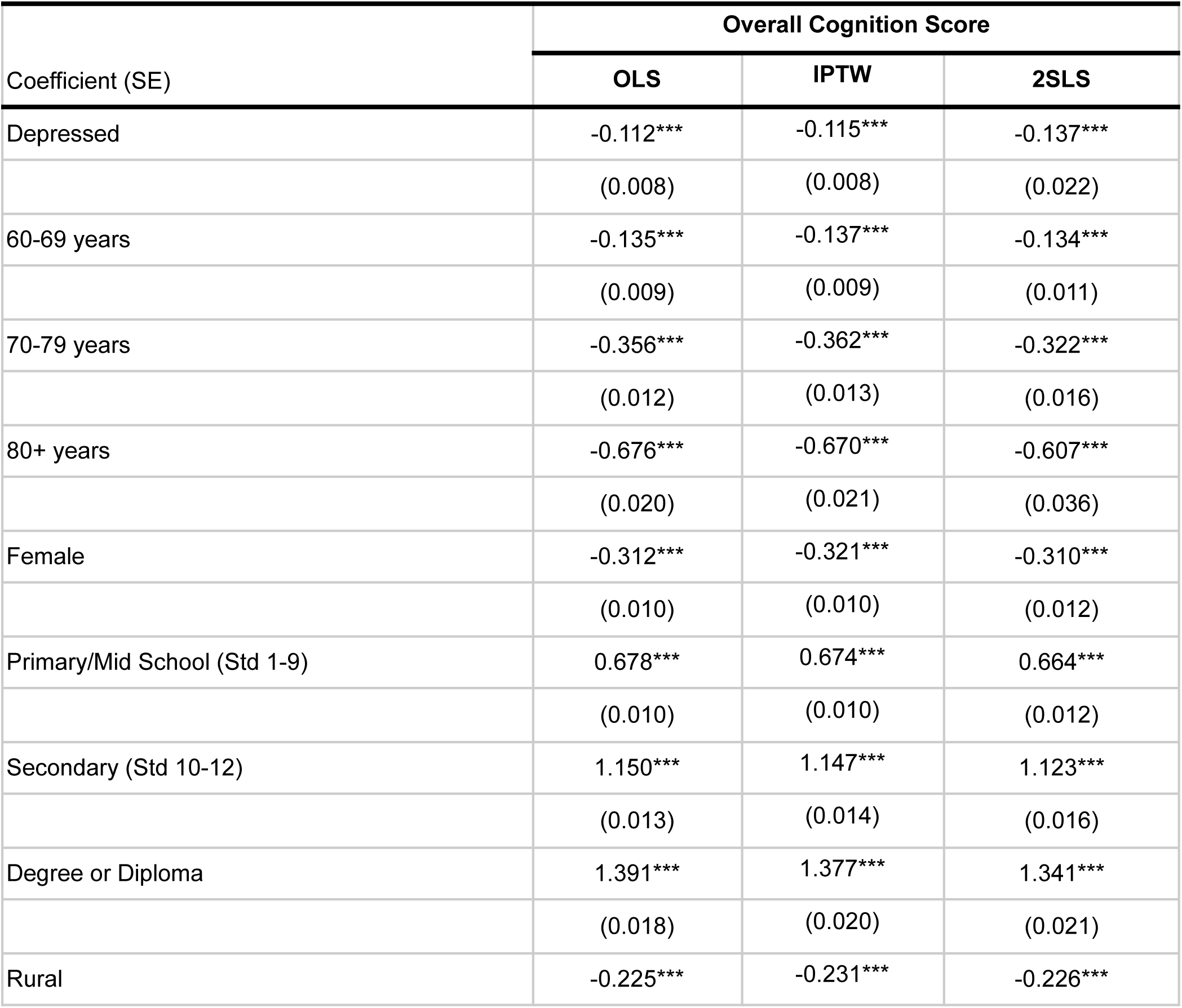
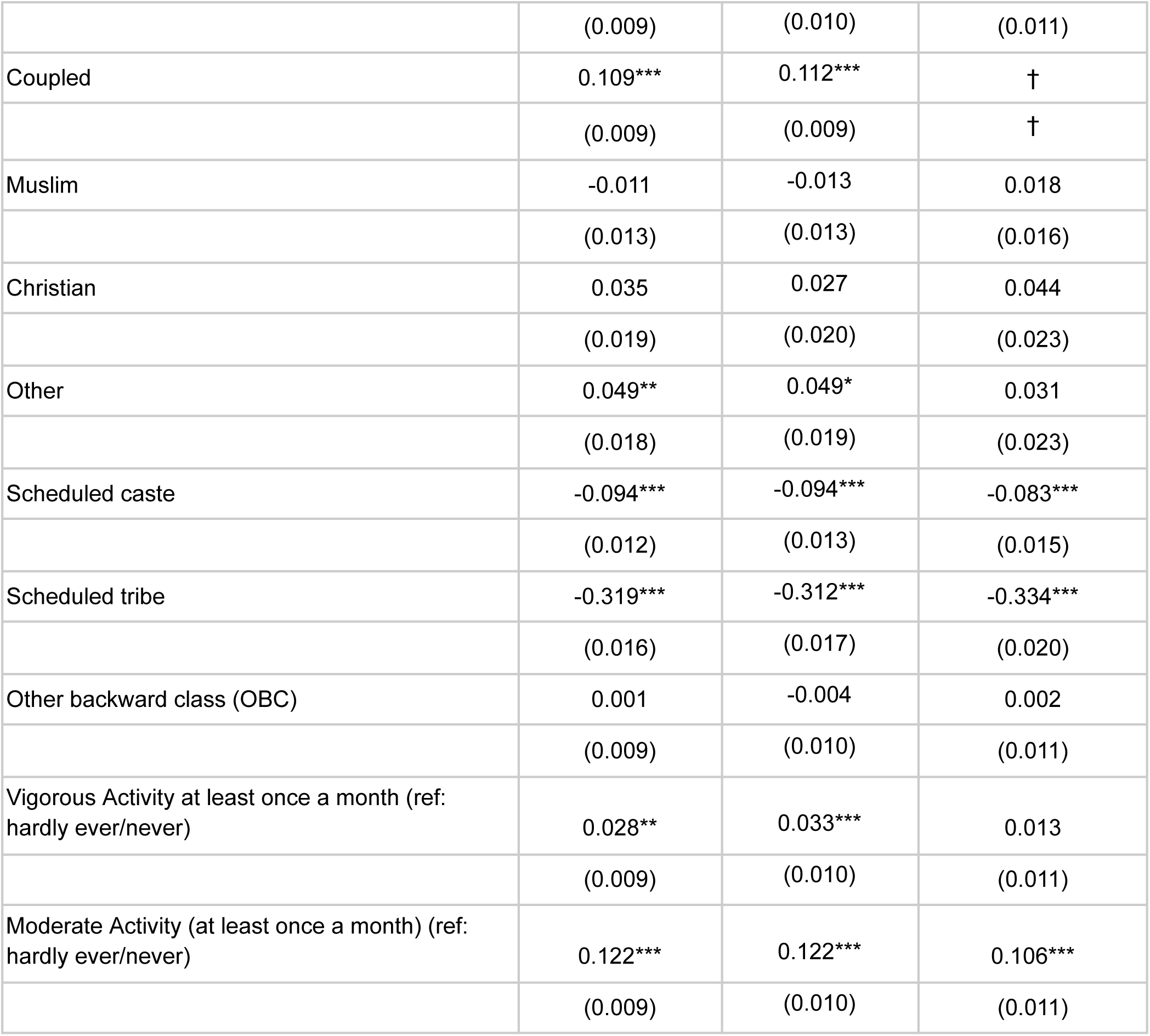
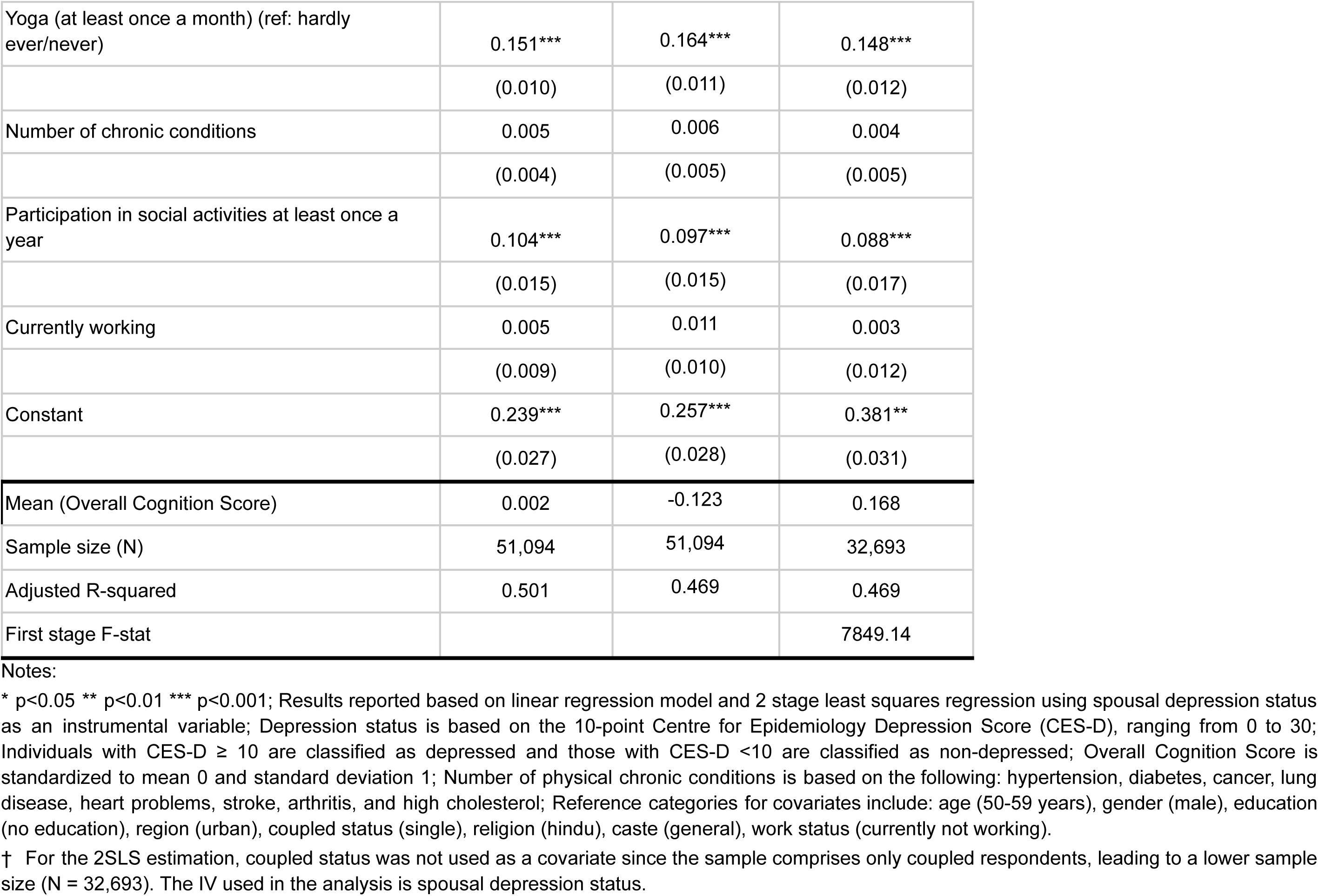
Association of Depression with Overall Cognition Score.

| Coefficient (SE) | Overall Cognition Score |  |  |
| --- | --- | --- | --- |
|  | OLS | IPTW | 2SLS |
| Depressed | -0.112***<br>(0.008) | -0.115***<br>(0.008) | -0.137***<br>(0.022) |
| 60-69 years | -0.135***<br>(0.009) | -0.137***<br>(0.009) | -0.134***<br>(0.011) |
| 70-79 years | -0.356***<br>(0.012) | -0.362***<br>(0.013) | -0.322***<br>(0.016) |
| 80+ years | -0.676***<br>(0.020) | -0.670***<br>(0.021) | -0.607***<br>(0.036) |
| Female | -0.312***<br>(0.010) | -0.321***<br>(0.010) | -0.310***<br>(0.012) |
| Primary/Mid School (Std 1-9) | 0.678***<br>(0.010) | 0.674***<br>(0.010) | 0.664***<br>(0.012) |
| Secondary (Std 10-12) | 1.150***<br>(0.013) | 1.147***<br>(0.014) | 1.123***<br>(0.016) |
| Degree or Diploma | 1.391***<br>(0.018) | 1.377***<br>(0.020) | 1.341***<br>(0.021) |
| Rural | -0.225*** | -0.231*** | -0.226*** |
|  | (0.009) | (0.010) | (0.011) |
| Coupled | 0.109*** | 0.112*** | † |
|  | (0.009) | (0.009) | † |
| Muslim | -0.011 | -0.013 | 0.018 |
|  | (0.013) | (0.013) | (0.016) |
| Christian | 0.035 | 0.027 | 0.044 |
|  | (0.019) | (0.020) | (0.023) |
| Other | 0.049** | 0.049* | 0.031 |
|  | (0.018) | (0.019) | (0.023) |
| Scheduled caste | -0.094*** | -0.094*** | -0.083*** |
|  | (0.012) | (0.013) | (0.015) |
| Scheduled tribe | -0.319*** | -0.312*** | -0.334*** |
|  | (0.016) | (0.017) | (0.020) |
| Other backward class (OBC) | 0.001 | -0.004 | 0.002 |
|  | (0.009) | (0.010) | (0.011) |
| Vigorous Activity at least once a month (ref: hardly ever/never) | 0.028** | 0.033*** | 0.013 |
|  | (0.009) | (0.010) | (0.011) |
| Moderate Activity (at least once a month) (ref: hardly ever/never) | 0.122*** | 0.122*** | 0.106*** |
|  | (0.009) | (0.010) | (0.011) |
| Yoga (at least once a month) (ref: hardly ever/never) | 0.151*** | 0.164*** | 0.148*** |
|  | (0.010) | (0.011) | (0.012) |
| Number of chronic conditions | 0.005 | 0.006 | 0.004 |
|  | (0.004) | (0.005) | (0.005) |
| Participation in social activities at least once a year | 0.104*** | 0.097*** | 0.088*** |
|  | (0.015) | (0.015) | (0.017) |
| Currently working | 0.005 | 0.011 | 0.003 |
|  | (0.009) | (0.010) | (0.012) |
| Constant | 0.239*** | 0.257*** | 0.381** |
|  | (0.027) | (0.028) | (0.031) |
| Mean (Overall Cognition Score) | 0.002 | -0.123 | 0.168 |
| Sample size (N) | 51,094 | 51,094 | 32,693 |
| Adjusted R-squared | 0.501 | 0.469 | 0.469 |
| First stage F-stat |  |  | 7849.14 |

Notes:
\* $p < 0.05$ \*\* $p < 0.01$ \*\*\* $p < 0.001$ ; Results reported based on linear regression model and 2 stage least squares regression using spousal depression status as an instrumental variable; Depression status is based on the 10-point Centre for Epidemiology Depression Score (CES-D), ranging from 0 to 30; Individuals with CES-D $\geq 10$ are classified as depressed and those with CES-D $< 10$ are classified as non-depressed; Overall Cognition Score is standardized to mean 0 and standard deviation 1; Number of physical chronic conditions is based on the following: hypertension, diabetes, cancer, lung disease, heart problems, stroke, arthritis, and high cholesterol; Reference categories for covariates include: age (50-59 years), gender (male), education (no education), region (urban), coupled status (single), religion (hindu), caste (general), work status (currently not working).
† For the 2SLS estimation, coupled status was not used as a covariate since the sample comprises only coupled respondents, leading to a lower sample size (N = 32,693). The IV used in the analysis is spousal depression status.

The similarity in propensity score distribution between depressed and non-depressed groups (Figure A2) supports the use of the IPTW regression model. The standardized mean differences in covariate values before and after IPTW are shown in the Appendix (Figure A3). After matching, the standardized mean difference was within ±10%, indicating an effective matching.

Table 4 reports the results of the association of depression status with each domain of cognitive function. We noted the strongest association for the domains of language (OLS: b=-0.146, p<0.001; 2SLS: b=-0.282, p<0.001) and immediate memory (OLS: b=-0.100, p<0.001; 2SLS: b=-0.134, p<0.001), and weakest for visuospatial ability (OLS: b=-0.026, p<0.01). The results from IPTW regression are reported in the Appendix (Table A4).

**Table 4.** Association of Depression with Cognition Domain Scores.

| **Panel A** | **Orientation** |  | **Immediate Memory** |  | **Delayed Memory** |  |
| --- | --- | --- | --- | --- | --- | --- |
| Coefficient (SE) | **OLS** | **2SLS** | **OLS** | **2SLS** | **OLS** | **2SLS** |
| Depression | -0.091*** | -0.103*** | -0.100*** | -0.134*** | -0.079*** | -0.082** |
|  | (0.009) | (0.025) | (0.010) | (0.029) | (0.010) | (0.030) |
| Sample Size (N) | 51,094 | 32,693 | 51,094 | 32,693 | 51,094 | 32,693 |
| Adjusted R-squared | 0.325 | 0.296 | 0.167 | 0.139 | 0.134 | 0.112 |
| First stage F-stat |  | 7849.14 |  | 7849.14 |  | 7849.14 |
| Mean (Cognition Score) | 0.002 | 0.153 | 0.002 | 0.105 | 0.000 | 0.085 |
| **Panel B** | **Language** |  | **Executive Function** |  | **Visuospatial Ability** |  |
| Coefficient (SE) | **OLS** | **2SLS** | **OLS** | **2SLS** | **OLS** | **2SLS** |
| Depression | -0.146*** | -0.282*** | -0.060*** | -0.049* | -0.026** | -0.029 |
|  | (0.015) | (0.041) | (0.009) | (0.025) | (0.008) | (0.025) |
| Sample Size (N) | 24,812 | 17,718 | 51,094 | 32,693 | 51,094 | 32,693 |
| Adjusted R-squared | 0.17 | 0.143 | 0.396 | 0.367 | 0.406 | 0.394 |
| First stage F-stat |  | 3882.77 |  | 7849.14 |  | 7849.14 |
| Mean (Cognition Score) | 0.008 | 0.052 | 0.000 | 0.149 | -0.001 | 0.118 |

Notes:
\* $p < 0.05$ \*\* $p < 0.01$ \*\*\* $p < 0.001$ ; Results reported based on linear regression model and 2 stage least squares regression using spousal depression status as an instrumental variable; Depression status is based on the 10-point Centre for Epidemiology Depression Score (CES-D), ranging from 0 to 30. Individuals with CES-D $\geq 10$ are classified as depressed and those with CES-D $< 10$ are classified as non-depressed; Each Cognition Domain Score is standardized to mean 0 and standard deviation 1; Covariates included in the model include: age, gender, education, region, coupled status, religion, caste, vigorous physical activity, moderate physical activity, yoga/meditation, yearly social activities, and work status.

### 3.3 Mediation Analysis

The results of the mediation analyses are reported in Figure 4 and Table 5. Figure 4 shows the diagrammatic representations of the three mediator variables and the extent to which they account for the association between depression and cognitive function. We noted depression to be significantly associated with higher ADL/IADL disability scores and lower quality of sleep (path *a* in Figure 4). Moreover, higher ADL/IADL disability score (better sleep quality score) was associated with significantly lower (higher) cognition score (path *b* in Figure 4). The proportion of the total effect mediated was highest for IADL (26.29%), followed by ADL (15.45%), and sleep quality (10.78%) (see Table 5).

**Figure 4.**
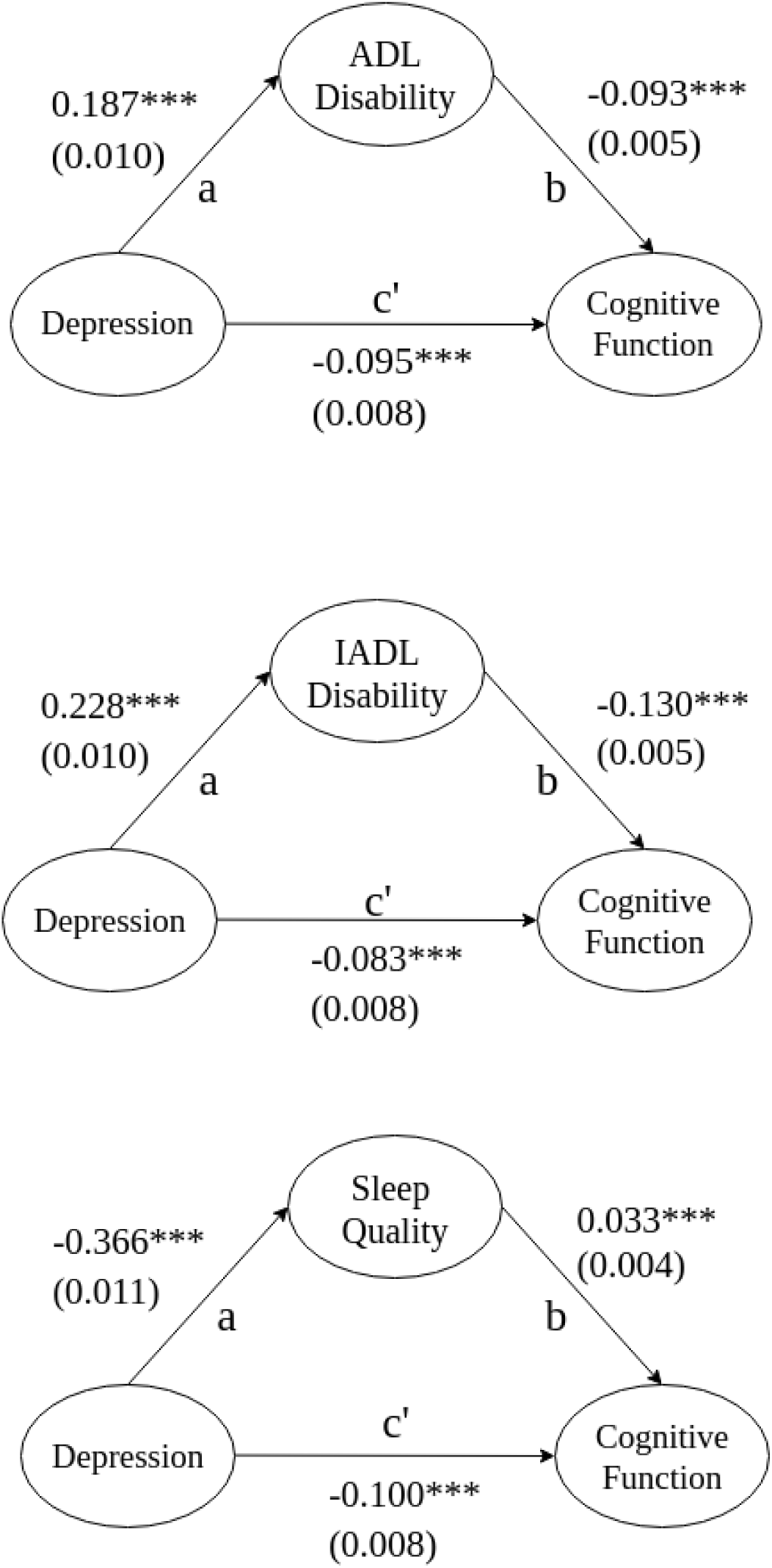
Mediation of the Association of Depression with Overall Cognitive Function. Notes: * p<0.05 ** p<0.01 *** p<0.001; The Natural Direct Effect (NDE) is indicated by path *c*’, the Natural Indirect Effect (NIE) is the product of the effect of the Treatment on the Mediator (*a*) and the effect of the Mediator on the Outcome (*b*) and denoted by *ab*. The total effect (TE) of the Treatment on the Outcome is the sum of NDE and NIE (*c*’ + *ab*).

**Table 5.** Mediation Analysis: Pathways from Depression to Cognitive Function.

| Coefficient (SE) | ADL | IADL | Sleep Quality |
| --- | --- | --- | --- |
| Natural Indirect Effect (NIE) | -0.017*** | -0.029*** | -0.012*** |
|  | (0.001) | (0.002) | (0.002) |
| Natural Direct Effect (NDE) | -0.095*** | -0.082*** | -0.100*** |
|  | (0.008) | (0.008) | (0.008) |
| Total Effect (TE) | -0.112*** | -0.112*** | -0.112*** |
|  | (0.008) | (0.008) | (0.008) |
| Proportion Mediated (NIE/TE) | 15.45% | 26.29% | 10.78% |

Notes:
\* $p < 0.05$ \*\* $p < 0.01$ \*\*\* $p < 0.001$ . The Natural Indirect Effect (NIE) is calculated as the product of the coefficient of treatment variable in mediation equation ( $a$ in Figure 4) with the coefficient of the mediator in the outcome equation ( $b$ in Figure 4); The Natural Direct Effect (NDE) is defined as the coefficient of the treatment variable in the outcome equation ( $c'$ in Figure 4) and Total Effect (TE) is the sum of NIE and NDE; The proportion mediated, calculated as $NIE/TE$ denotes the percentage of the total effect operating through the indirect channel.

### 3.4 Heterogeneity Analysis

We also assessed the heterogeneity in the association between depression status and cognitive function by socio-economic and demographic factors: age, gender, education, region, and coupled status (Figure 5). The association of depression with cognitive function score was strongest for the 80+ age group; moreover, the association was significantly higher for the 80+ age cohort compared to those in the 50-59 and 60-69 age cohorts.

**Figure 5.**
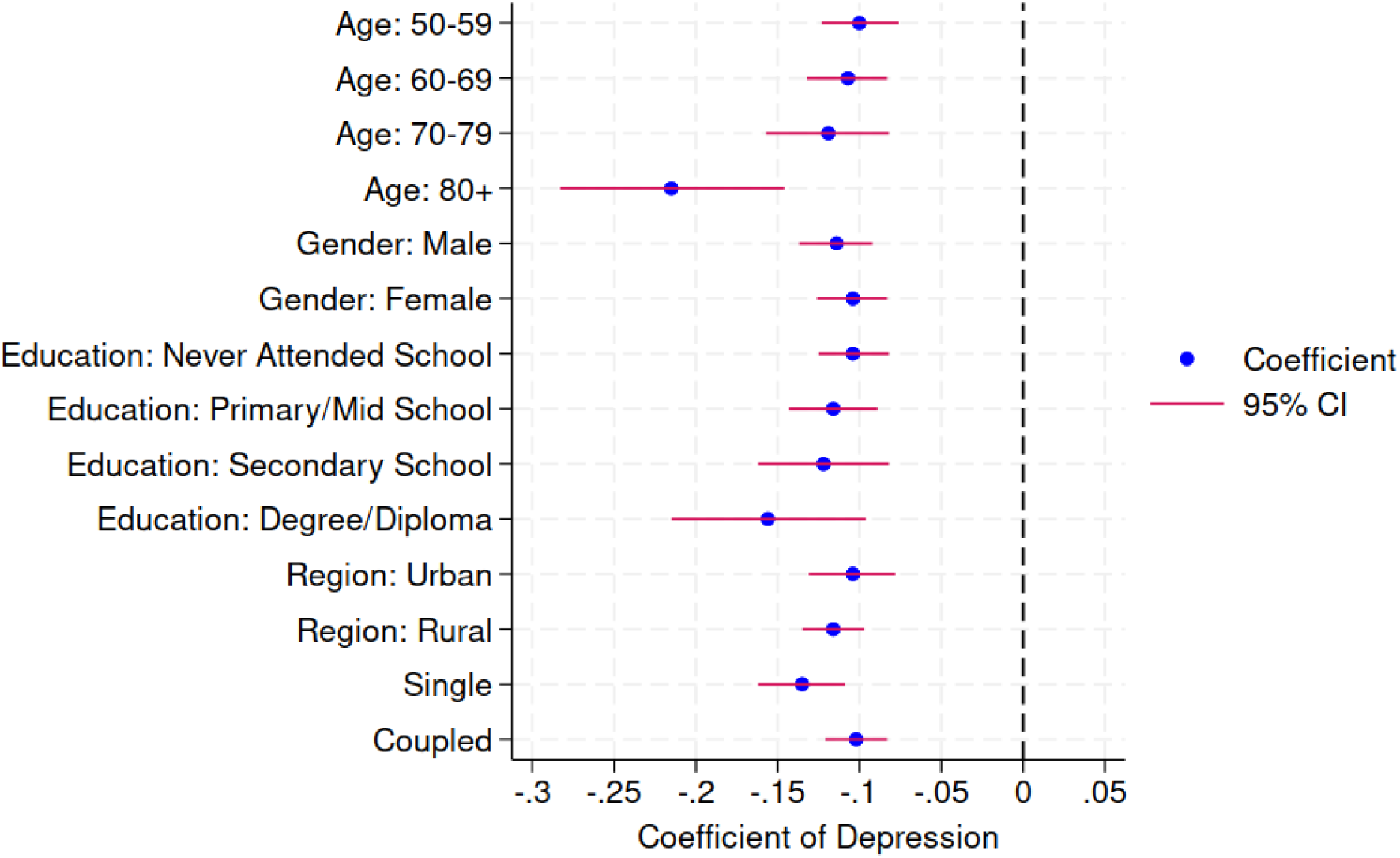
Heterogeneity Analysis: Association of Depression with Overall Cognitive Function. Notes: The plotted values (x-axis) indicate the coefficient of depression (dichotomous measure based on a cutoff of 10 or more in the CES-D score) in the regression model with standardized overall cognition factor score as the outcome variable, when each of the categories defined by the categorical variables (y-axis) are considered individually as the sample for the analysis.

Figure 6 depicts heterogeneity in the mediation effect by depressive symptom severity (CES-D score was categorized as 0-4, 5-9, 10-14, 14-19, and ≥20). The plots indicate that the NIE, NDE, and TE increase with the severity of depressive symptoms. The gradient of NIE is steeper for ADL/IADL compared to sleep quality indicating that the mediation effects of ADL/IADL are stronger at higher levels of depressive symptom severity compared to those for sleep quality.

**Figure 6.**
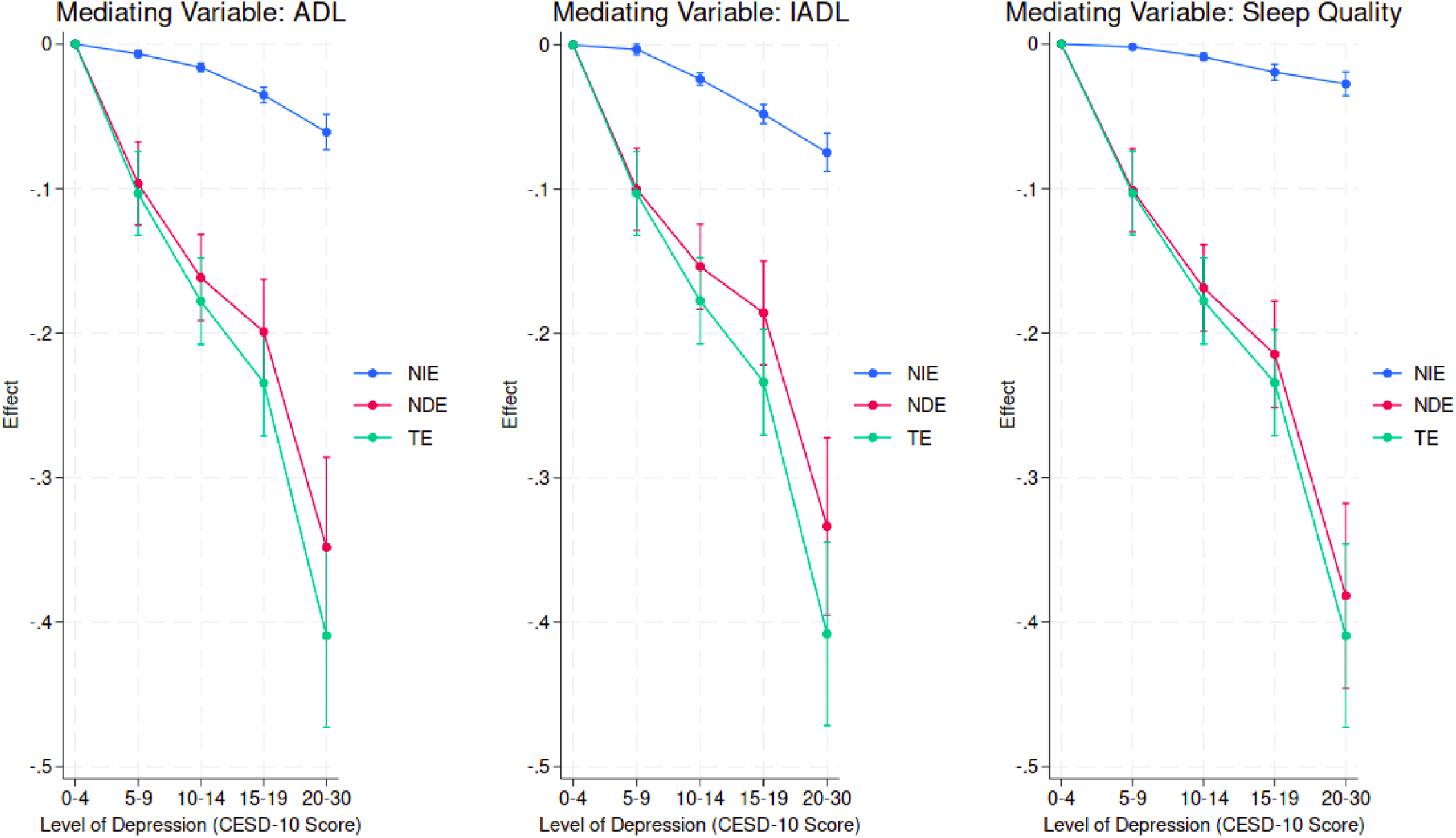
Heterogeneity Analysis: Mediation Analysis for Different Levels of Depressive Symptoms. Notes: For this analysis, the severity of depressive symptomatology has been categorized using an ordered categorical variable defined on the CES-D score with 5 levels: 0-4, 5-9, 10-14, 15-19, 20-30; Higher values indicate more severe depressive symptoms; The Natural Indirect Effect (NIE) is calculated as the product of the coefficient of treatment variable in mediation equation (*a* in Figure 4) with the coefficient of the mediator in the outcome equation (*b* in Figure 4); The Natural Direct Effect (NDE) is defined as the coefficient of the treatment variable in the outcome equation (*c*’ in Figure 4) and Total Effect (TE) is the sum of NIE and NDE; Line plots indicate the trends in NIE, NDE and TE with the severity of depression, with higher negative magnitudes indicating higher effects.

### 3.5 Robustness Analysis

We performed additional analysis to assess the robustness of the results to an alternative measure for the overall cognition score. We defined a binary indicator (0/1) identifying those in the bottom quartile of overall cognition factor score as cognitively impaired. Based on this measure, we found individuals in the depressed group to have 3.2% higher risk of cognitive impairment (Linear Probability Model: Risk ratio=1.032, p<0.001; 2SLS: Risk ratio=1.026, p<0.05) (see Appendix Table A5).

Additionally, we performed robustness analysis using alternative (higher) cutoff values for depression symptomatology – 12 and 15 – instead of 10. We noted that the proportion of individuals meeting criteria for depressive symptoms using the cutoffs of 12 and 15 reduced to 27.8% and 12.0%, respectively. The results are reported in the Appendix (Table A6). We consistently found significant association of depressive symptoms with overall cognition score, although the magnitude of the association was amplified using the higher cutoffs of 12 (OLS: b=-0.120, p<0.001; 2SLS: b= −0.142, p<0.001) and 15 (OLS: b=-0.140, p<0.001; 2SLS: b=-0.182, p<0.001).

## 4. Discussion

In this study, we examined the association of depressive symptoms on cognitive performance and explored the socio-economic and demographic correlates of depression and cognition among middle-aged and elderly adults in India using data from LASI Wave I, 2017-18. Previous studies have explored this association in various populations; our research builds on this by examining the possible causal impact of depression on cognitive function and investigating selected mediators. To our knowledge, no prior study has examined this relationship in the Indian context. Using alternative estimation methods – inverse probability of treatment weighting and instrumental variable regression – to account for potential selection on observed and unobserved determinants of depression status we assessed their plausible causal relationship. We also explored potential pathways and estimated the direct and indirect effects linking depression with cognitive function by using ADL, IADL, and sleep quality as mediators. Our results highlighted the debilitating effect of depressive symptoms on several domains of cognitive function (viz. language, memory, orientation, and executive function). We also noted significant mediation effects, driven by IADL (26.3%), ADL (15.5%), and sleep problems (10.8%).

We compared the deficits observed across several cognitive domains associated with depressive symptoms. Notably, language and immediate memory domains were most significantly affected. This is consistent with the findings of Kreische et al. (2022), who noted strong differences between depressed patients and healthy controls in verbal learning and memory, verbal fluency and working memory. We hypothesized substantial reduction in executive function associated with depressive symptoms; however, the effects were smaller compared to those for language and memory domains. One possible explanation is that the included tasks in executive functioning do not fully capture the extent of cognitive functionality involved, such as planning and analytical thinking. From a neurological perspective, depression causes atrophy of important brain regions such as hippocampus (associated with memory), insula (associated with socio-emotional processing), and STG (associated with language), which lead to social withdrawal, communication problems, and a lack of relatedness with the world.

We also explored the role of sleep quality, ADL, and IADL in mediating the association between depression and cognitive performance. Several studies have discussed the effects of depression on different regions of the brain and neurochemistry, which in turn lead to cognitive deficits (Zackova et al., 2021; Butters et al., 2008; Sawyer et al., 2012; Diniz et al., 2013; Xu et al., 2024). These effects can manifest as lifestyle changes such as social withdrawal, poor sleep quality, communication problems, and functional disabilities, which can lead to worsening of cognitive health. Recent work has explored the impact of sleep on cognitive performance among individuals with mood disorders (Pearson et al., 2023). Sleep quality can mediate the association between cognitive impairment and depression (Liu et al., 2022; Liu and Chen, 2024; Wu et al., 2019; Guo et al., 2021). Sleep problems (insomnia/hypersomnia) are considered a core symptom of depression and have a bi-directional causal link with the disorder (Riemann et al., 2001). Moreover, sleep has well-established associations with cognitive decline (Torossian et al., 2021). The CES-D 20-point scale (Radloff, 1977) includes a question on sleep quality, whereas the 10-point version used in LASI does not.

ADL and IADL disabilities have a bi-directional relationship with depressive symptoms (Zhou et al., 2024, Wang et al., 2023). Fan et al. (2024) developed a chain mediation model to describe the mediating role of social activities and ADL in the path between MDD and cognitive impairment, using the data of China Health and Retirement Longitudinal Study. Depression has been positively associated with loss of ADL capabilities in large ageing populations in China (Liu et al., 2023, Zhou et al., 2024, Wang et al., 2023, Zhao et al., 2022). Peng et al. (2023) discuss the impact of depression and ADL disability on cognitive performance. IADL disability has also proven to be a significant mediator in the pathway between depression and cognitive impairment (Liu et al., 2023). Our findings indicate a substantially higher proportion of the total effect of depression on cognitive function being mediated by IADL (26.29%) and ADL (15.45%). Depression affects the brain regions responsible for these functions and is associated with symptoms like lack of motivation, apathy, and anhedonia (Pizzagalli, 2014). These symptoms can make performing ADL/IADL tasks feel less meaningful, leading to poorer performance, which in turn may contribute to cognitive decline.

We recognize several limitations of our study. Despite the alternative estimation approaches (IPTW and IV regression models) to account for observed and unobserved confounding, we cannot rule out the presence of residual confounding. Prior work suggests that the cognitive domains such as processing speed, attention, and inhibition are strongly affected by depression (Kriesche et al., 2022; Ahern et al., 2024; Rock et al., 2014). However, since LASI does not contain information aimed at capturing these domains, we are limited in our understanding of how depression affects these domains of cognition. In our mediation analysis, although we have explored ADL, IADL, and sleep quality as mediators, we cannot rule out the possibility of other mediators which can lie in the pathway between depression and cognition. Finally, as noted earlier, there may exist a bi-directional relationship between the mediator and the treatment variable (e.g. ADL/IADL disability leading to depression) and between the mediator and outcome variable (e.g. cognitive declining resulting in limitations in ADL/IADLs), which we were unable to capture in our analysis due to lack of suitable IVs - one for depression and one for the mediator.

In summary, our research provides robust evidence that depression is significantly associated with diminished cognitive performance in middle-aged and older adults. This relationship is further influenced by mediating factors such as limitations in activities of daily living (ADL), instrumental activities of daily living (IADL), and sleep quality. Utilizing data from India, our study extends the applicability of these findings to low- and middle-income countries characterized by diverse socio-demographic profiles and varying healthcare infrastructures. These results underscore the urgent need for targeted interventions addressing mental health and cognitive decline in aging populations to promote overall well-being in later life.

## Data Availability

All data used in the study are publicly available.

https://g2aging.org/lasi/download

## Acknowledgement

This analysis utilized data from the Harmonized LASI dataset and Codebook, Version A.3 as of April 2023, developed by the Gateway to Global Aging Data (DOI: https://doi.org/10.25549/h-lasi). The development of the Harmonized LASI was funded by the National Institute on Aging (R01 AG042778, 2R01 AG030153, 2R01 AG051125). For more information about the Harmonization project, please refer to https://g2aging.org/.

## Appendix

**Figure A1:**
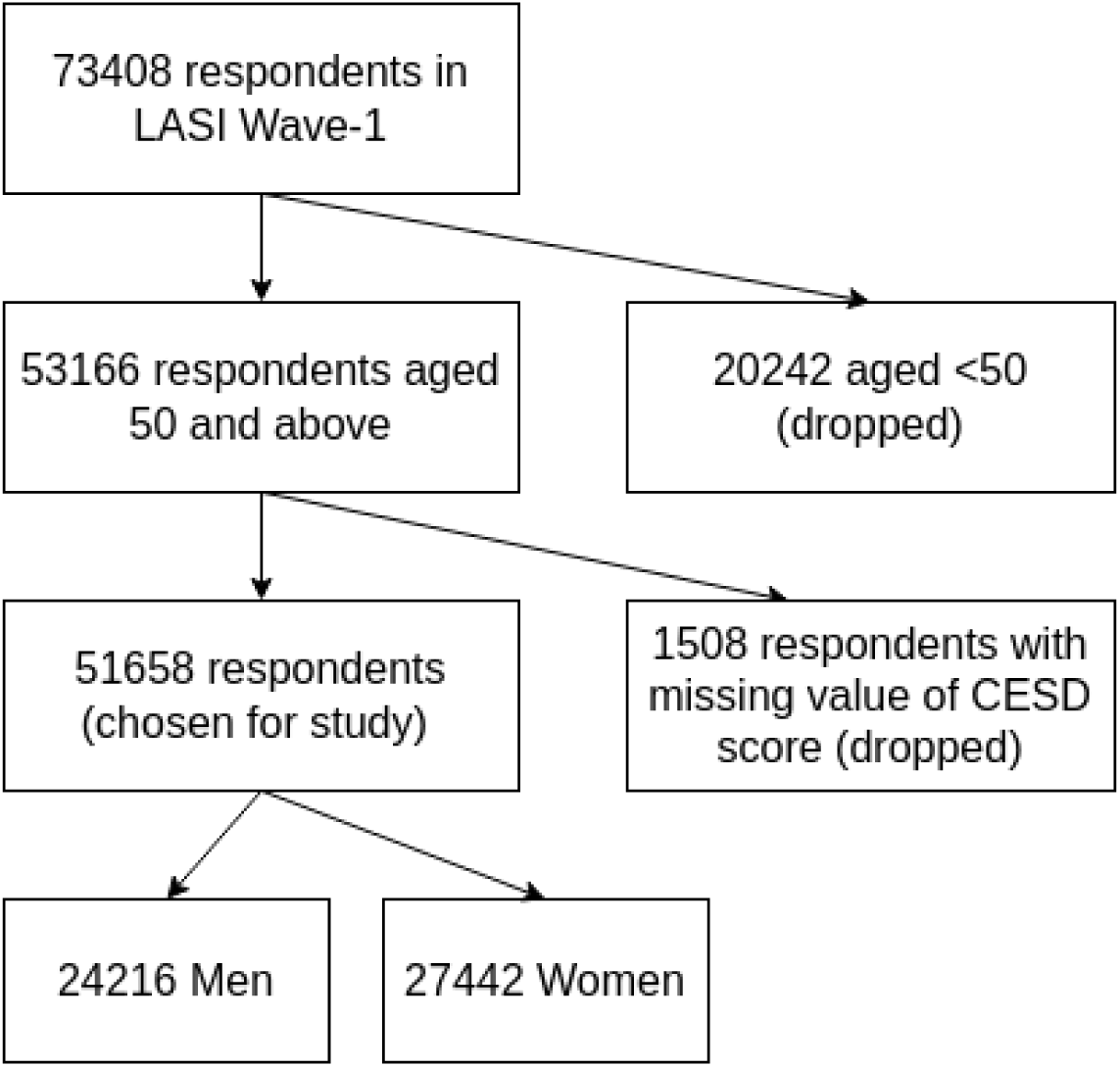
Study sample selection criteria.

**Table A1:**
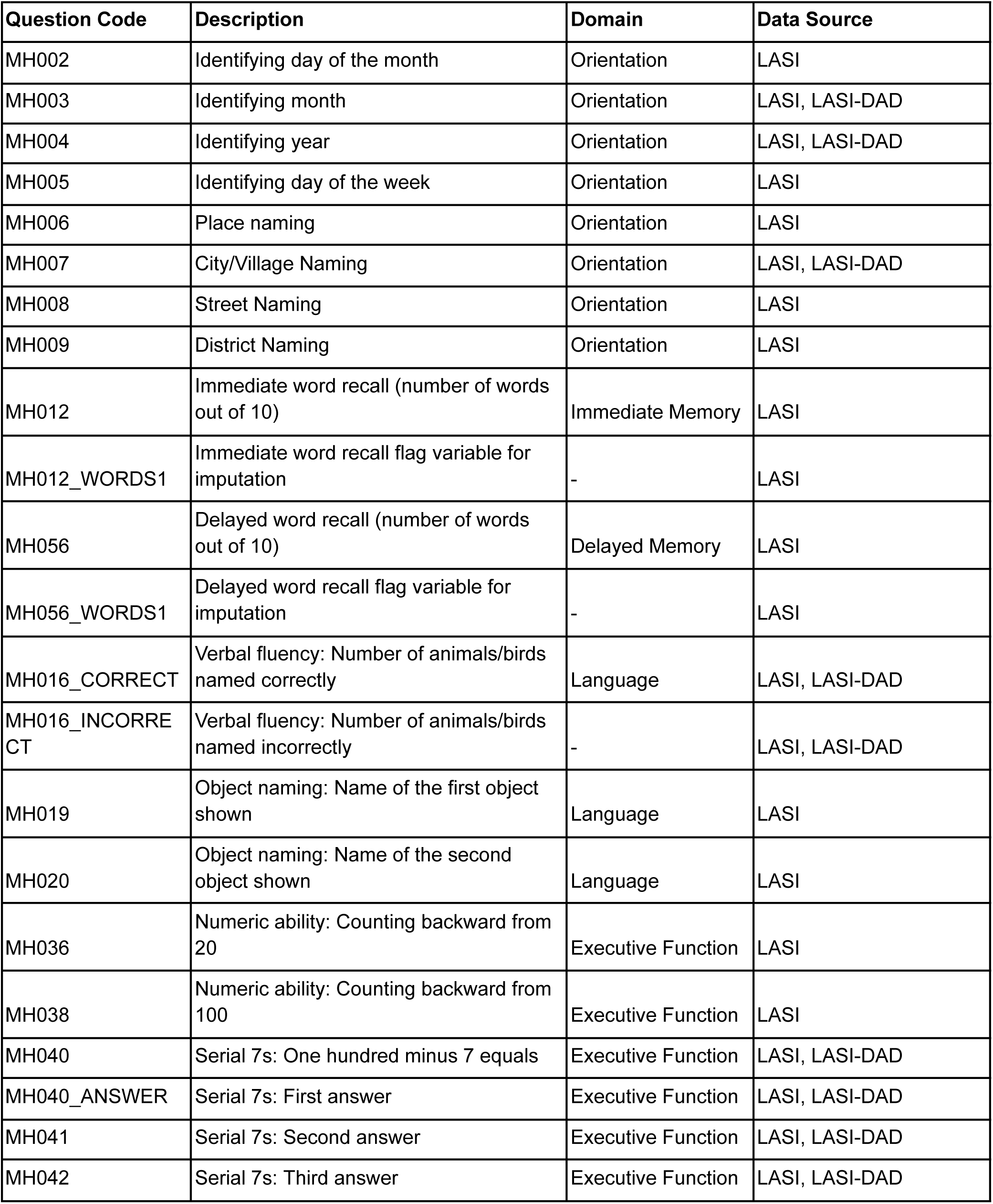
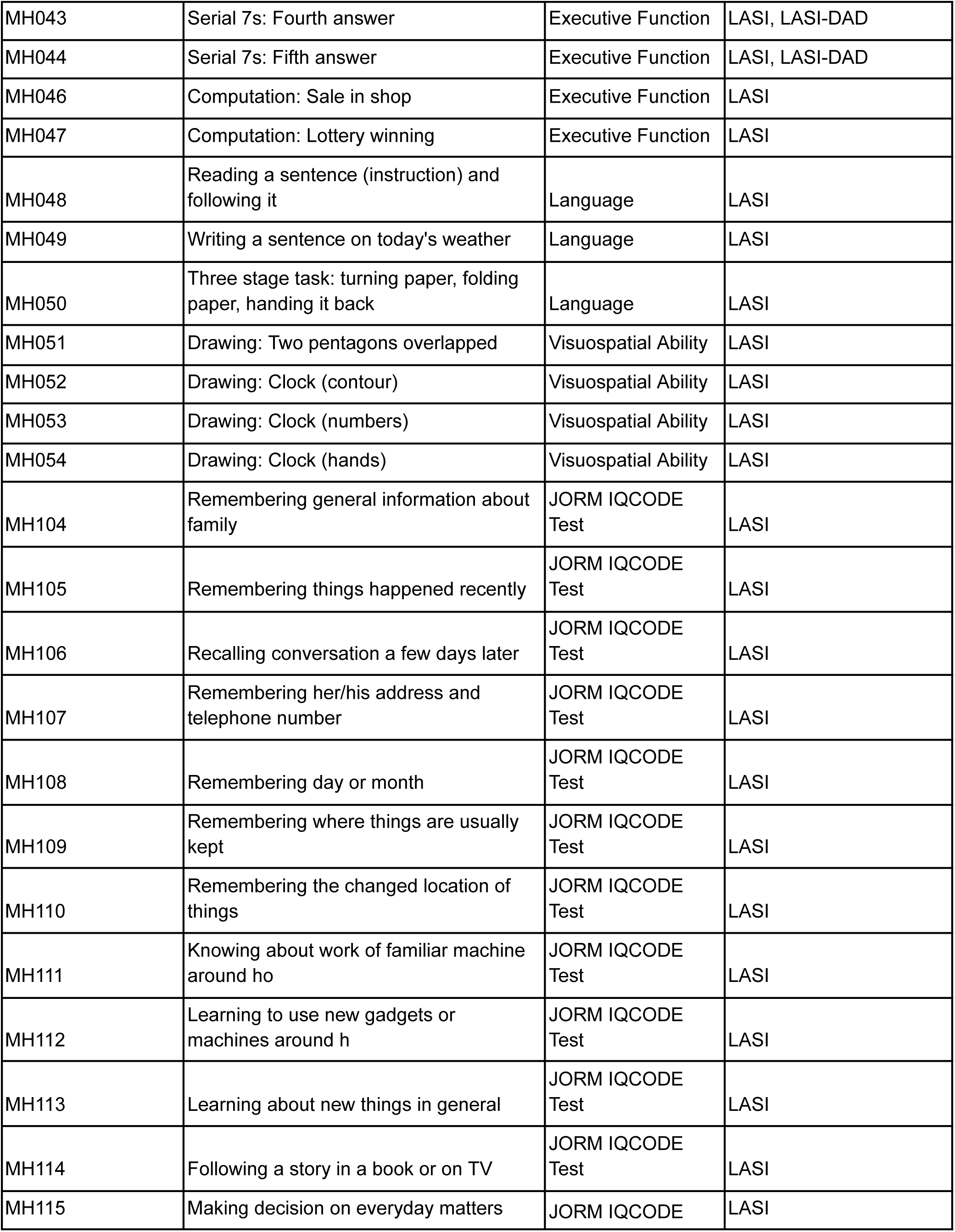
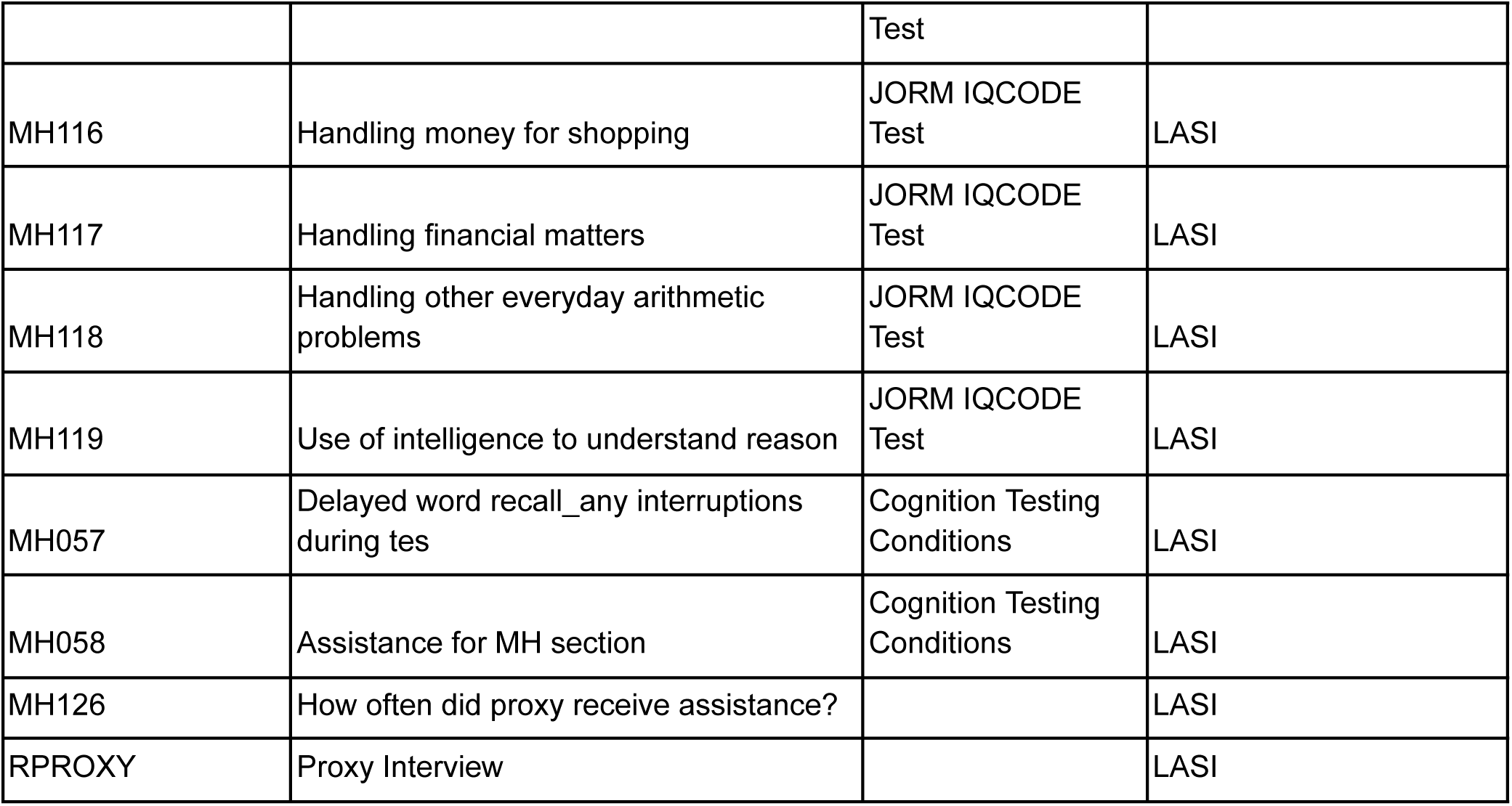
Items included in overall factor score.

**Table A2:** Items included in each Cognitive Domain.

| Variable name | Variable Label | Range | Cognitive Domain |
| --- | --- | --- | --- |
| r1mo | Identifying current month | 0-1 | Orientation |
| r1dy | Identifying day of the month | 0-1 |  |
| r1dw | Identifying day of the week | 0-1 |  |
| r1yr | Identifying current year | 0-1 |  |
| r1place | Place naming | 0-1 |  |
| r1city | City naming | 0-1 |  |
| r1address | Street naming | 0-1 |  |
| r1dist | District naming | 0-1 |  |
| r1imrc | 10-word immediate recall | 0-10 | Immediate Memory |
| r1dlrc | 10-word delayed recall | 0-10 | Delayed Memory |
| r1verbf | Verbal fluency (naming animals) | 0-61 | Language |
| r1read | Reading a sentence | 0-1 |  |
| r1senten | Writing a sentence | 0-1 |  |
| r1execu | 3-stage task | 0-3 |  |
| r1object | Object naming (2 objects asked; number of correct answers) | 0-2 |  |
| r1bwc100a | Backward counting from 100 | 0-1 | Executive function |
| r1bwc20a | Backward counting from 20 | 0-1 |  |
| r1ser7 | Serial 7s (Number of correct subtractions) | 0-5 |  |
| r1compu | Computations (2 questions asked; number of correct answers) | 0-2 |  |
| r1draw | Drawing interlocked pentagons | 0-1 | Visuospatial |
| r1drawcl | Drawing a clock | 0-3 |  |

Notes:
This system of classification has been borrowed from the appendix of the following reference: Nichols, E., Gross, A.L., Hu, P. et al. The association between BMI and cognition in India: data from the Longitudinal Aging Study in India (LASI). BMC Public Health 24, 2720 (2024). https://doi.org/10.1186/s12889-024-20101-y. Only the domain of “Memory” has been split into Immediate Memory and Delayed memory by us.

**Table A3:** Weighted Summary Statistics by Depression Status with Non-Standardized Values for Cognition Scores and Mediators.

|  | **Depressed** | **Non-depressed** | **Total** | **p-value\*** |
| --- | --- | --- | --- | --- |
|  | (N=22,691) | (N=28,967) | (N=51,658) |  |
| Cognition Scores |  |  |  |  |
| Overall Cognition, Mean (SD) | -0.183 (0.957) | 0.119 (0.975) | -0.020 (0.978) | <0.001 |
| Orientation, Mean (SD) | 0.815 (0.193) | 0.860 (0.174) | 0.840 (0.185) | <0.001 |
| Immediate Memory, Mean (SD) | 4.727 (1.762) | 5.111 (1.824) | 4.934 (1.806) | <0.001 |
| Delayed Memory, Mean (SD) | 3.627 (1.776) | 3.952 (1.914) | 3.802 (1.859) | <0.001 |
| Language, Mean (SD) | 0.081 (0.542) | 0.206 (0.514) | 0.156 (0.529) | <0.001 |
| Executive Function, Mean (SD) | -0.079 (0.762) | 0.095 (0.753) | 0.015 (0.762) | <0.001 |
| Visuospatial Ability, Mean (SD) | -0.159 (0.828) | 0.015 (0.887) | -0.065 (0.864) | <0.001 |
| Activities of Daily Living (ADL), Mean (SD) | 0.521 (1.267) | 0.249 (0.850) | 0.375 (1.071) | <0.001 |
| Instrumental Activities of Daily Living (IADL), Mean (SD) | 1.503 (2.120) | 0.854 (1.644) | 1.153 (1.906) | <0.001 |
| Sleep Quality, Mean (SD) | 2.629 (0.522) | 2.814 (0.393) | 2.729 (0.467) | <0.001 |

\* *p*-value based on two-sample *t*-test for continuous variables are reported; Depression status is based on the 10-point Centre for Epidemiology Depression Score (CES-D), ranging from 0 to 30. Individuals with CES-D $\geq 10$ are classified as depressed and those with CES-D $<10$ are classified as non-depressed.

**Figure A2:**
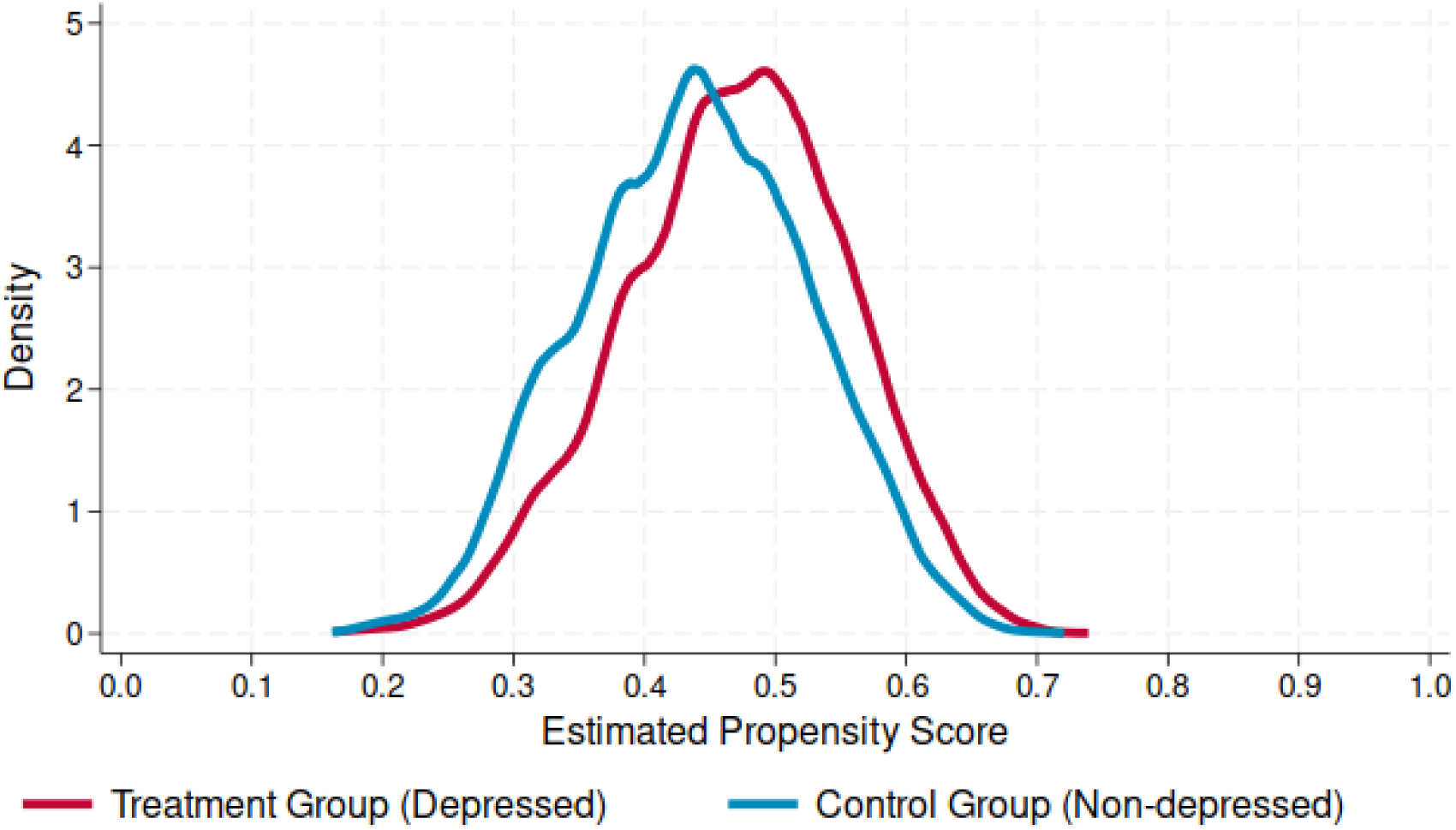
Overlap plot showing distributions of estimated propensity scores in IPTW before weighting. Notes: The above plot shows the kernel density estimates for observations in treatment (depressed) and control (non-depressed) groups before weighting done by IPTW. A good overlap between the treatment and control distributions ensures the validity of performing IPTW. The Epanechnikov kernel with a calculated optimal bandwidth has been used to generate the distributions.

**Figure A3:**
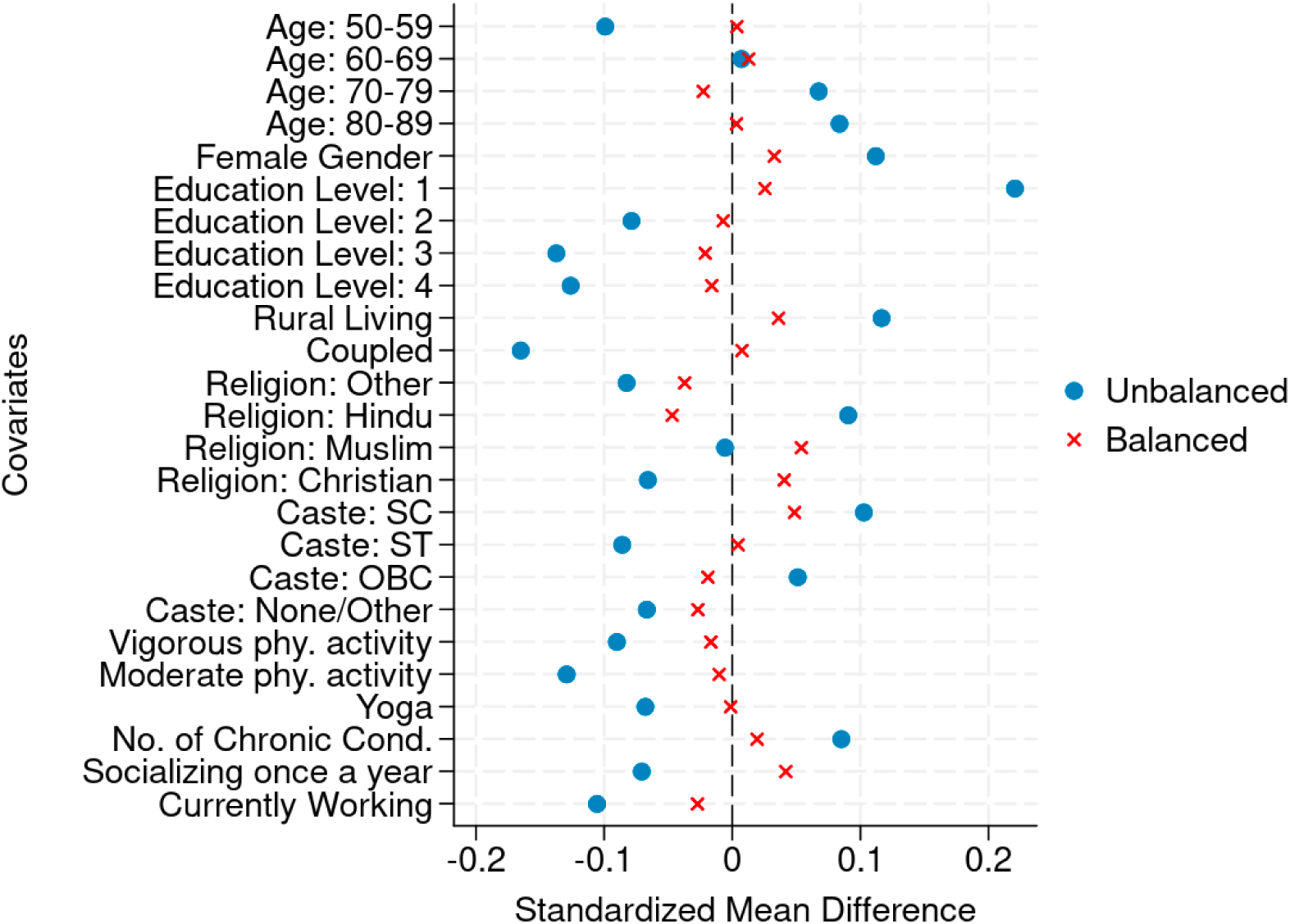
Love Plot from IPTW Notes: The above plot shows standardized mean difference in each of the covariates (difference in means between treatment and control groups standardized by the pooled standard deviation) before and after the weighting done by IPTW. Variables having more than two categories (namely age group, education level, religion and caste) are one-hot encoded into dummy variables. Number of chronic conditions is a continuous variable ranging from 0 to 7. All other variables are dichotomous. The balanced sample shows mean differences less than ±0.1 which indicates good matching done through IPTW.

**Table A4.**
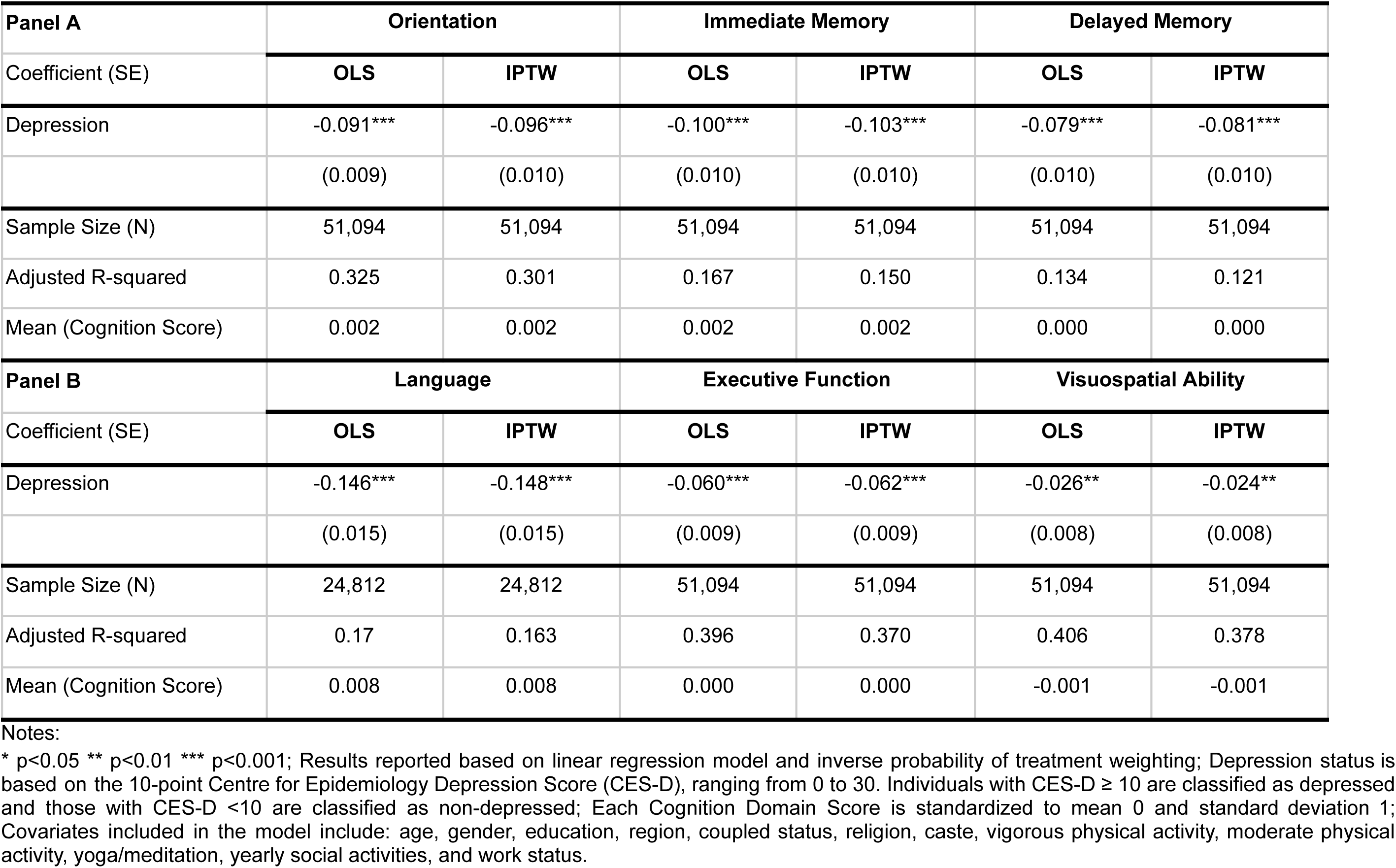
Association of Depression with Cognition Domain Scores (IPTW analysis)

| **Panel A** | **Orientation** |  | **Immediate Memory** |  | **Delayed Memory** |  |
| --- | --- | --- | --- | --- | --- | --- |
| Coefficient (SE) | **OLS** | **IPTW** | **OLS** | **IPTW** | **OLS** | **IPTW** |
| Depression | -0.091*** | -0.096*** | -0.100*** | -0.103*** | -0.079*** | -0.081*** |
|  | (0.009) | (0.010) | (0.010) | (0.010) | (0.010) | (0.010) |
| Sample Size (N) | 51,094 | 51,094 | 51,094 | 51,094 | 51,094 | 51,094 |
| Adjusted R-squared | 0.325 | 0.301 | 0.167 | 0.150 | 0.134 | 0.121 |
| Mean (Cognition Score) | 0.002 | 0.002 | 0.002 | 0.002 | 0.000 | 0.000 |
| **Panel B** | **Language** |  | **Executive Function** |  | **Visuospatial Ability** |  |
| Coefficient (SE) | **OLS** | **IPTW** | **OLS** | **IPTW** | **OLS** | **IPTW** |
| Depression | -0.146*** | -0.148*** | -0.060*** | -0.062*** | -0.026** | -0.024** |
|  | (0.015) | (0.015) | (0.009) | (0.009) | (0.008) | (0.008) |
| Sample Size (N) | 24,812 | 24,812 | 51,094 | 51,094 | 51,094 | 51,094 |
| Adjusted R-squared | 0.17 | 0.163 | 0.396 | 0.370 | 0.406 | 0.378 |
| Mean (Cognition Score) | 0.008 | 0.008 | 0.000 | 0.000 | -0.001 | -0.001 |

Notes:
\* $p < 0.05$ \*\* $p < 0.01$ \*\*\* $p < 0.001$ ; Results reported based on linear regression model and inverse probability of treatment weighting; Depression status is based on the 10-point Centre for Epidemiology Depression Score (CES-D), ranging from 0 to 30. Individuals with CES-D $\geq 10$ are classified as depressed and those with CES-D $< 10$ are classified as non-depressed; Each Cognition Domain Score is standardized to mean 0 and standard deviation 1; Covariates included in the model include: age, gender, education, region, coupled status, religion, caste, vigorous physical activity, moderate physical activity, yoga/meditation, yearly social activities, and work status.

**Table A5.** Robustness Analysis: Association of Depression with Dichotomous measure of Cognitive Impairment.

| Odds ratio (SE) | Cognitive Impairment |  |
| --- | --- | --- |
|  | LPM | 2SLS |
| Depressed | 1.032*** | 1.026* |
|  | (0.004) | (0.012) |
| 60-69 years | 1.051*** | 1.048*** |
|  | (0.005) | (0.006) |
| 70-79 years | 1.158*** | 1.125*** |
|  | (0.008) | (0.010) |
| 80+ years | 1.332*** | 1.282*** |
|  | (0.015) | (0.025) |
| Female | 1.150*** | 1.140*** |
|  | (0.006) | (0.008) |
| Primary/Mid School (Std 1-9) | 0.794*** | 0.813*** |
|  | (0.004) | (0.005) |
| Secondary (Std 10-12) | 0.778*** | 0.798*** |
|  | (0.004) | (0.005) |
| Degree or Diploma | 0.795*** | 0.813*** |
|  | (0.005) | (0.006) |
| Rural | 1.085*** | 1.071*** |
|  | (0.005) | (0.005) |
| Coupled | 0.948*** | - |
|  | (0.005) | - |
| Muslim | 1.003 | 0.988 |
|  | (0.007) | (0.008) |
| Christian | 0.999 | 0.982 |
|  | (0.010) | (0.012) |
| Other | 0.984 | 0.988 |
|  | (0.010) | (0.011) |
| Scheduled caste | 1.045*** | 1.037*** |
|  | (0.007) | (0.008) |
| Scheduled tribe | 1.153*** | 1.160*** |
|  | (0.011) | (0.014) |
| Other backward class (OBC) | 1.010* | 1.011* |
|  | (0.005) | (0.006) |
| Vigorous Activity at least once a month (ref: hardly ever/never) | 0.988** | 0.995 |
|  | (0.005) | (0.006) |
| Moderate Activity (at least once a month) (ref: hardly ever/never) | 0.957*** | 0.969*** |
|  | (0.005) | (0.006) |
| Yoga (at least once a month) (ref: hardly ever/never) | 0.947*** | 0.950*** |
|  | -0.005 | (0.005) |
| Number of chronic conditions | 0.991*** | 0.991*** |
|  | (0.002) | (0.003) |
| Participation in social activities at least once a year | 0.985* | 0.992 |
|  | (0.007) | (0.008) |
| Currently working | 1.006 | 1.006 |
|  | (-0.005) | (0.006) |
| Constant | 1.087*** | 1.036*** |
|  | (0.016) | (0.018) |
| Sample size (N) | 51,094 | 32,693 |
| R-squared | 0.246 | 0.194 |
| First stage F-stat |  | 7849.14 |
| Mean (Cognitive Impairment) | 0.250 | 0.188 |

Notes:
\* $p < 0.05$ \*\* $p < 0.01$ \*\*\* $p < 0.001$ ; LPM: Linear Probability Model; Cognitive impairment is defined as a binary variable such that individuals who belong to the bottom 25% quantile of the standardized Overall Cognitive Score are defined as being cognitively impaired while the others are defined as the healthy control group; Results reported based on linear probability model and 2 stage least squares regression using spousal depression status as an instrumental variable; Odds ratios are reported along with standard errors in parenthesis; Depression status is based on the 10-point Centre for Epidemiology Depression Score (CES-D), ranging from 0 to 30. Individuals with CES-D $\geq 10$ are classified as depressed and those with CES-D $< 10$ are classified as non-depressed; Overall Cognition Score is standardized to mean 0 and standard deviation 1; number of physical chronic conditions is based on the following: hypertension, diabetes, cancer, lung disease, heart problems, stroke, arthritis, and high cholesterol; Reference categories for covariates include: age (50-59 years), gender (male), education (no education), region (urban), coupled status (single), religion (hindu), caste (general), work status (currently not working).

**Table A6.**
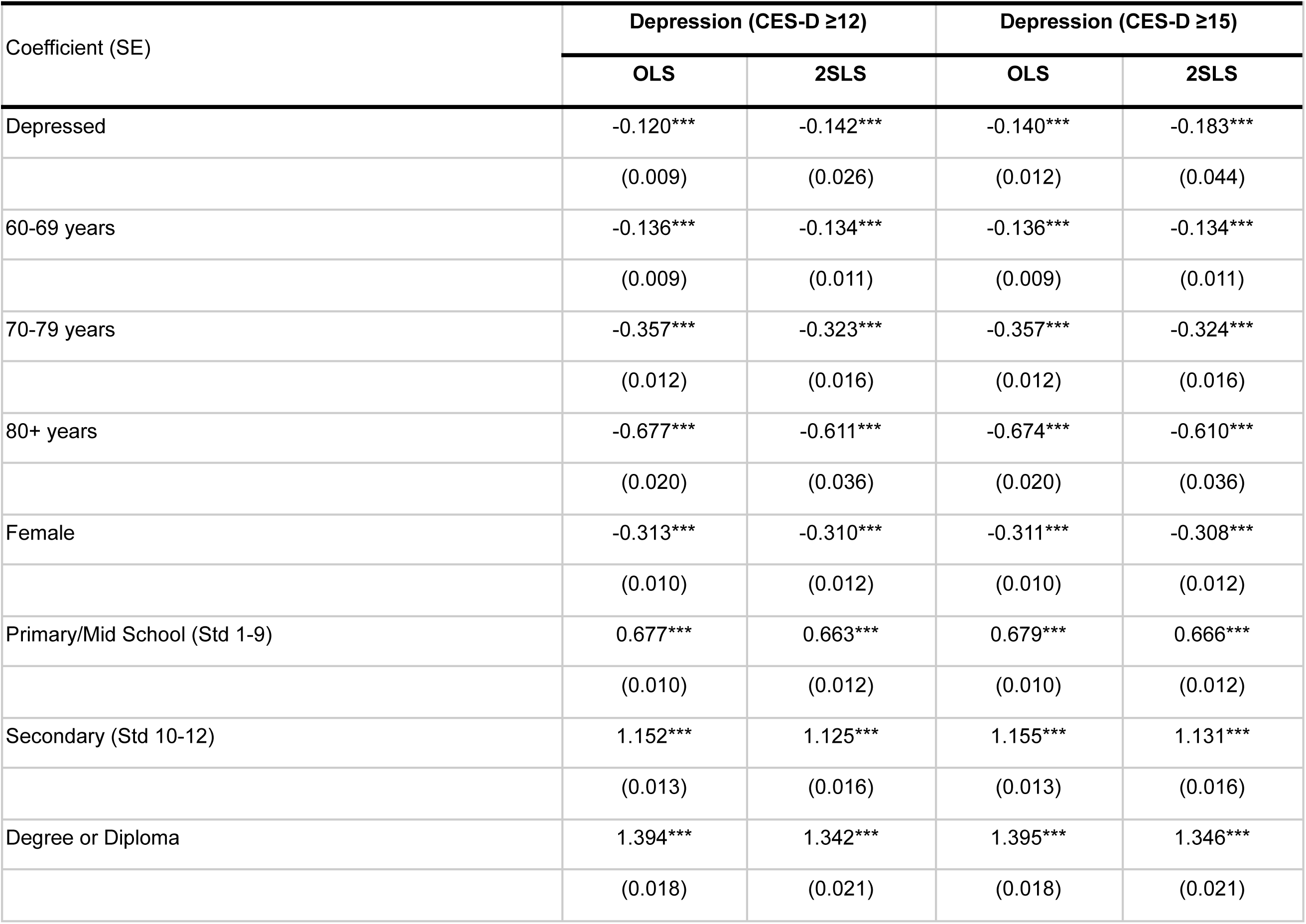
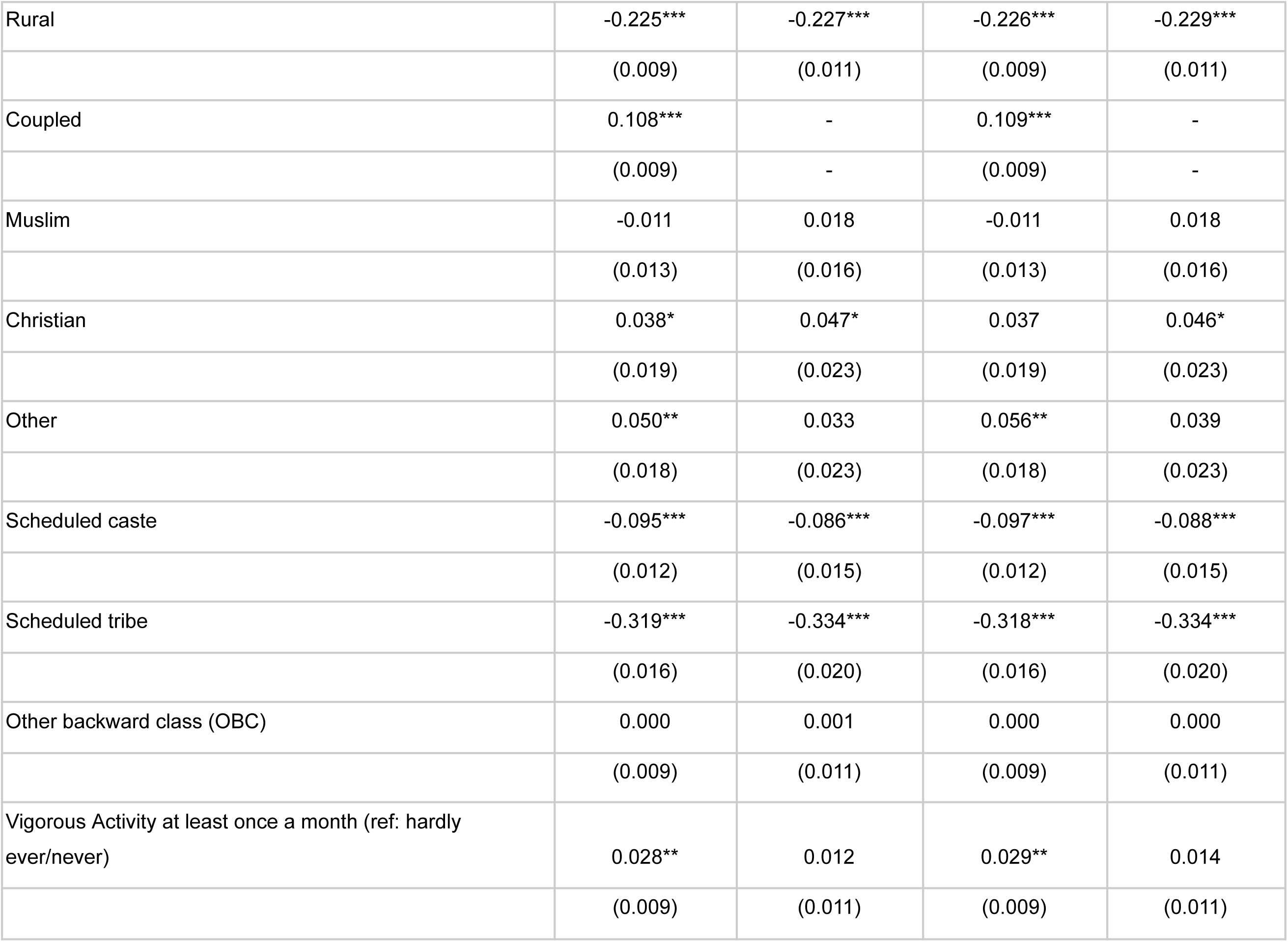
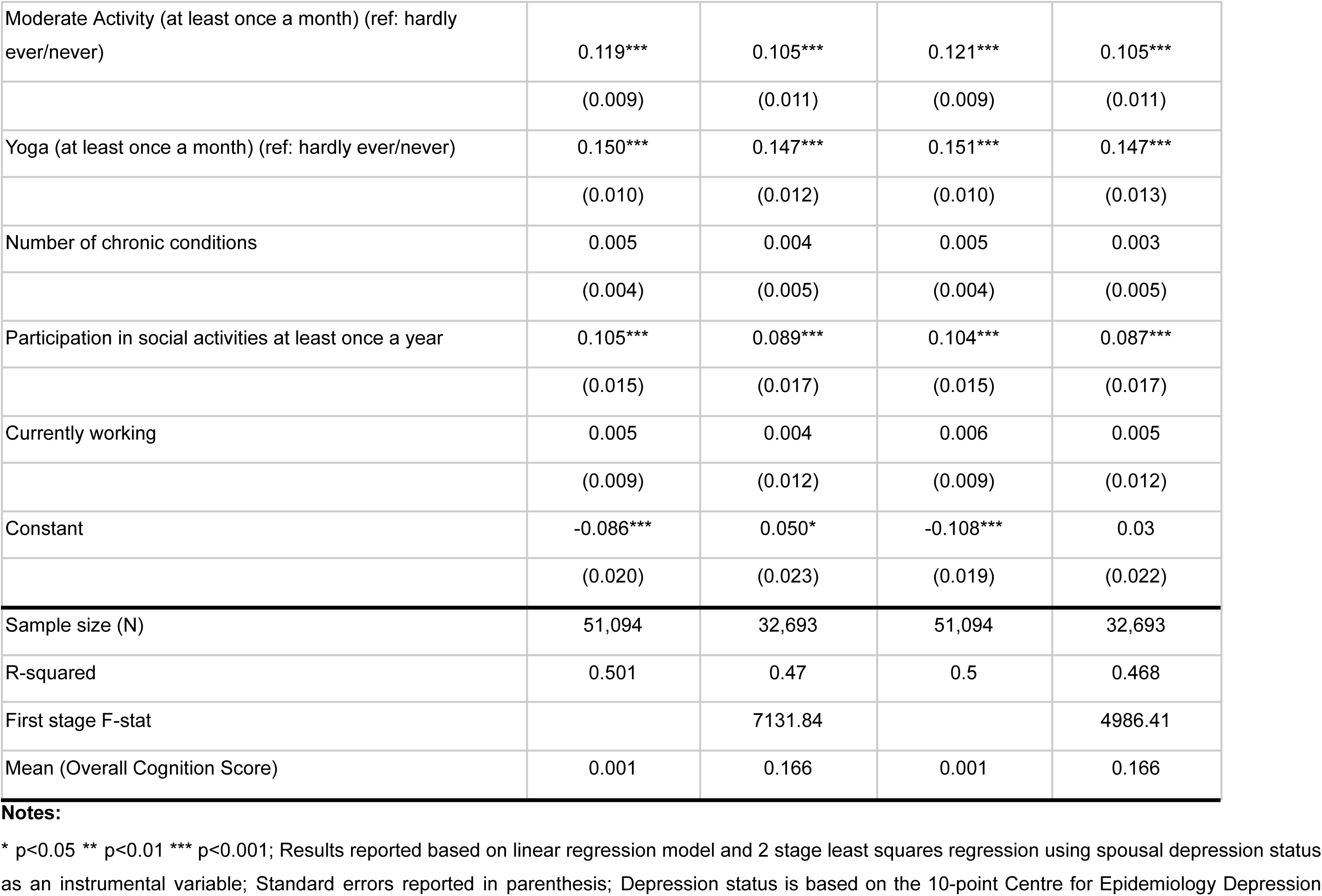
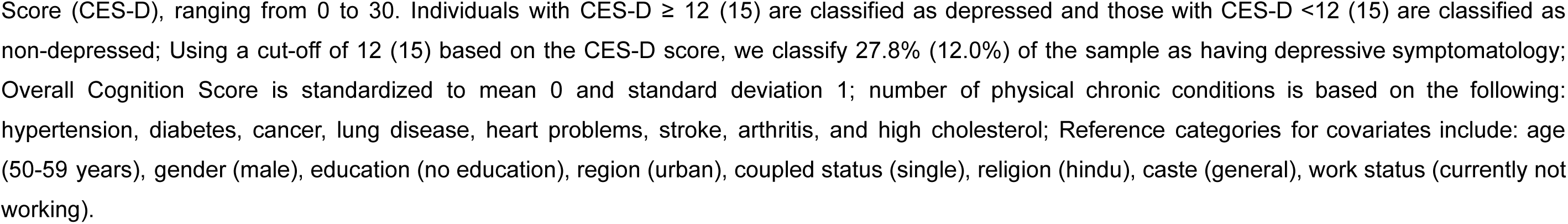
Robustness Analysis: Association of Depression with Overall Cognition Score Using Alternative Cut-off for Depression.

| Coefficient (SE) | Depression (CES-D $\geq 12$) | | Depression (CES-D $\geq 15$) | |
| --- | --- | --- | --- | --- |
|  | OLS | 2SLS | OLS | 2SLS |
| Depressed | -0.120*** | -0.142*** | -0.140*** | -0.183*** |
|  | (0.009) | (0.026) | (0.012) | (0.044) |
| 60-69 years | -0.136*** | -0.134*** | -0.136*** | -0.134*** |
|  | (0.009) | (0.011) | (0.009) | (0.011) |
| 70-79 years | -0.357*** | -0.323*** | -0.357*** | -0.324*** |
|  | (0.012) | (0.016) | (0.012) | (0.016) |
| 80+ years | -0.677*** | -0.611*** | -0.674*** | -0.610*** |
|  | (0.020) | (0.036) | (0.020) | (0.036) |
| Female | -0.313*** | -0.310*** | -0.311*** | -0.308*** |
|  | (0.010) | (0.012) | (0.010) | (0.012) |
| Primary/Mid School (Std 1-9) | 0.677*** | 0.663*** | 0.679*** | 0.666*** |
|  | (0.010) | (0.012) | (0.010) | (0.012) |
| Secondary (Std 10-12) | 1.152*** | 1.125*** | 1.155*** | 1.131*** |
|  | (0.013) | (0.016) | (0.013) | (0.016) |
| Degree or Diploma | 1.394*** | 1.342*** | 1.395*** | 1.346*** |
|  | (0.018) | (0.021) | (0.018) | (0.021) |
| Rural | -0.225*** | -0.227*** | -0.226*** | -0.229*** |
|  | (0.009) | (0.011) | (0.009) | (0.011) |
| Coupled | 0.108*** | - | 0.109*** | - |
|  | (0.009) | - | (0.009) | - |
| Muslim | -0.011 | 0.018 | -0.011 | 0.018 |
|  | (0.013) | (0.016) | (0.013) | (0.016) |
| Christian | 0.038* | 0.047* | 0.037 | 0.046* |
|  | (0.019) | (0.023) | (0.019) | (0.023) |
| Other | 0.050** | 0.033 | 0.056** | 0.039 |
|  | (0.018) | (0.023) | (0.018) | (0.023) |
| Scheduled caste | -0.095*** | -0.086*** | -0.097*** | -0.088*** |
|  | (0.012) | (0.015) | (0.012) | (0.015) |
| Scheduled tribe | -0.319*** | -0.334*** | -0.318*** | -0.334*** |
|  | (0.016) | (0.020) | (0.016) | (0.020) |
| Other backward class (OBC) | 0.000 | 0.001 | 0.000 | 0.000 |
|  | (0.009) | (0.011) | (0.009) | (0.011) |
| Vigorous Activity at least once a month (ref: hardly ever/never) | 0.028** | 0.012 | 0.029** | 0.014 |
|  | (0.009) | (0.011) | (0.009) | (0.011) |
| Moderate Activity (at least once a month) (ref: hardly ever/never) | 0.119*** | 0.105*** | 0.121*** | 0.105*** |
|  | (0.009) | (0.011) | (0.009) | (0.011) |
| Yoga (at least once a month) (ref: hardly ever/never) | 0.150*** | 0.147*** | 0.151*** | 0.147*** |
|  | (0.010) | (0.012) | (0.010) | (0.013) |
| Number of chronic conditions | 0.005 | 0.004 | 0.005 | 0.003 |
|  | (0.004) | (0.005) | (0.004) | (0.005) |
| Participation in social activities at least once a year | 0.105*** | 0.089*** | 0.104*** | 0.087*** |
|  | (0.015) | (0.017) | (0.015) | (0.017) |
| Currently working | 0.005 | 0.004 | 0.006 | 0.005 |
|  | (0.009) | (0.012) | (0.009) | (0.012) |
| Constant | -0.086*** | 0.050* | -0.108*** | 0.03 |
|  | (0.020) | (0.023) | (0.019) | (0.022) |
| Sample size (N) | 51,094 | 32,693 | 51,094 | 32,693 |
| R-squared | 0.501 | 0.47 | 0.5 | 0.468 |
| First stage F-stat |  | 7131.84 |  | 4986.41 |
| Mean (Overall Cognition Score) | 0.001 | 0.166 | 0.001 | 0.166 |

**Notes:**
\* p<0.05 \*\* p<0.01 \*\*\* p<0.001; Results reported based on linear regression model and 2 stage least squares regression using spousal depression status as an instrumental variable; Standard errors reported in parenthesis; Depression status is based on the 10-point Centre for Epidemiology Depression
Score (CES-D), ranging from 0 to 30. Individuals with CES-D $\geq 12$ (15) are classified as depressed and those with CES-D $<12$ (15) are classified as non-depressed; Using a cut-off of 12 (15) based on the CES-D score, we classify 27.8% (12.0%) of the sample as having depressive symptomatology; Overall Cognition Score is standardized to mean 0 and standard deviation 1; number of physical chronic conditions is based on the following: hypertension, diabetes, cancer, lung disease, heart problems, stroke, arthritis, and high cholesterol; Reference categories for covariates include: age (50-59 years), gender (male), education (no education), region (urban), coupled status (single), religion (hindu), caste (general), work status (currently not working).

